# Pathogenic variants in myoferlin (*MYOF)* cause arrhythmogenic cardiomyopathy

**DOI:** 10.64898/2026.09.21.26363590

**Authors:** Anna Kirillova, Metin Aytekin, Irene Chan, Amir I. Mina, Yucheng Shao, Almina Kirdar, David Y. Zhang, Nishita Kalepalli, William S. Girard, Yassmin Y. Al Aaraj, Chloe Reuter, Jeffery S. Annis, Yunshan Yue, Siyi Jiang, Rashmi J. Rao, S. Mehdi Nouraie, Joseph Park, Ying Tang, Christopher Flores, Michael D. Creager, Paul J. Kim, Evan L. Brittain, Stuart A. Scott, Janet R. Manning, Satoshi Okawa, Guy Salama, Haodi Wu, Manling Zhang, Carlos J. Camacho, Michael S. Gold, Victoria N. Parikh, Stephen Y. Chan

## Abstract

Arrhythmogenic cardiomyopathy (ACM) is an inherited disease characterized by fibrofatty remodeling and sudden cardiac death linked to desmosomal dysfunction. Yet, 40% of cases remain genetically unexplained, and non-desmosomal mutations offer insight into ACM pathophysiology. Myoferlin (MYOF), a calcium (Ca²⁺)-binding protein in cardiomyocytes, is linked to cardiomyopathy, but its mechanism remains unknown. Whole-exome sequencing identified a heterozygous *MYOF* missense variant (p.G1654S) in an ACM family; we subsequently found three unrelated p.G1654S cases. We hypothesized that MYOF deficiency promotes Ca²⁺ handling deficits and arrhythmogenesis in ACM. In patient iPSC-derived cardiomyocytes, MYOF p.G1654S protein dimerized with wild-type protein and underwent accelerated lysosomal degradation, showed disrupted Ca²⁺ handling, and displayed increased arrhythmia burden – all rescued by genomic correction. MYOF interacted with CaV1.2, and verapamil pharmacologically rescued arrhythmias. Heterozygous *Myof* knockout mice showed systolic dysfunction and fibrosis. These findings establish MYOF as a causative ACM gene and therapeutic target that mediates arrhythmogenesis by disrupting MYOF-CaV1.2.

**One Sentence Summary:** MYOF deficiency disrupts MYOF-CaV1.2 interaction and calcium handling, causing arrhythmogenic cardiomyopathy.

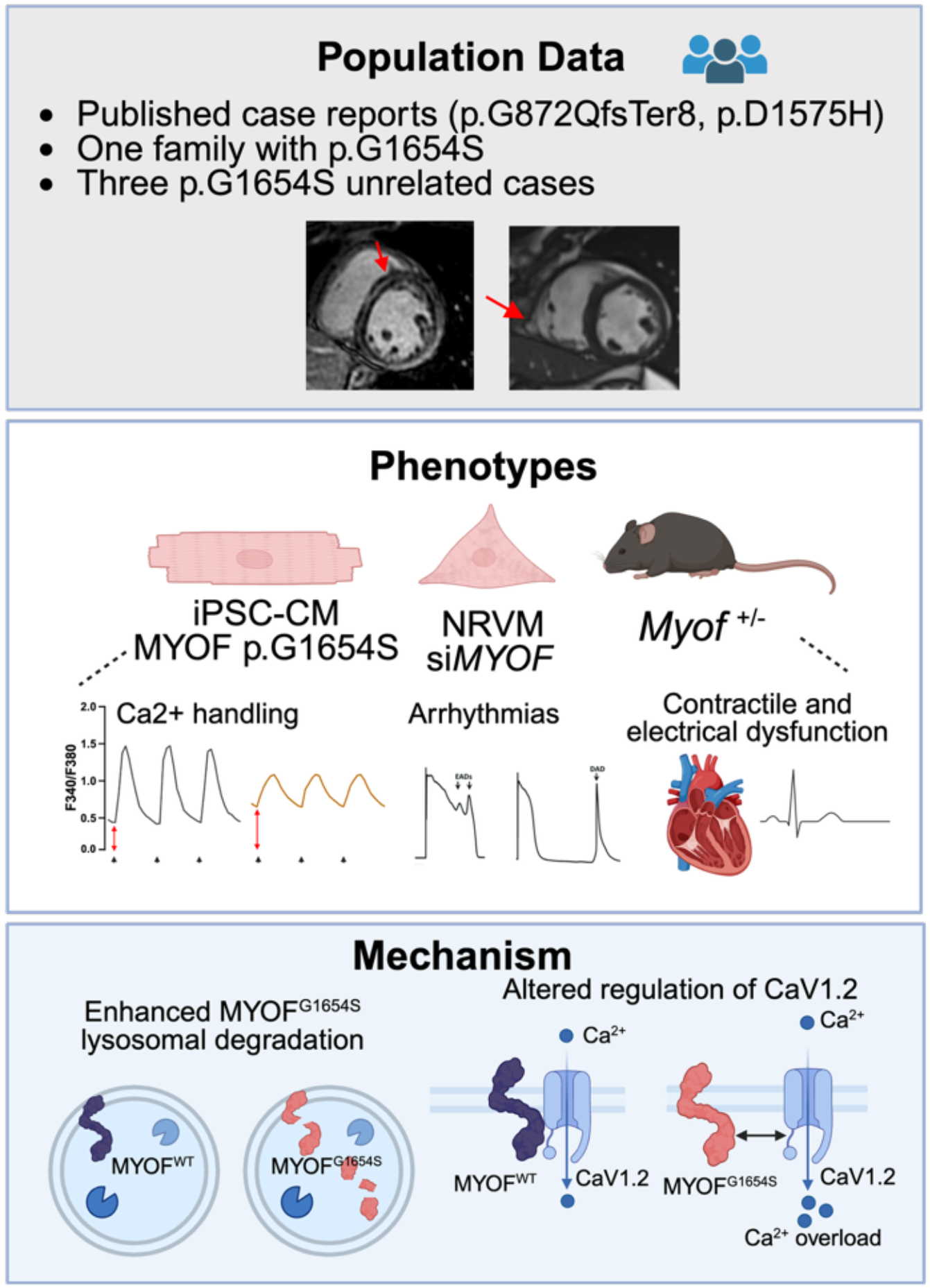

## Introduction

Inherited cardiomyopathies, such as arrhythmogenic right (or left) ventricular cardiomyopathy (ARVC; ALVC), can lead to sudden cardiac death and heart failure with reduced ejection fraction (HFrEF). Disruptions in desmosomal genes have been extensively described as significant causes of arrhythmogenic cardiomyopathy (ACM) pathogenesis (*1*). Yet, desmosome-based mechanisms do not fully explain arrhythmogenesis and ventricular contractile dysfunction in these cardiomyopathies. Non-desmosomal disruptions are increasingly appreciated as drivers of ACM, but their pathogenic mechanisms remain poorly understood (*2*). Furthermore, only ∼20% of ACM patients carry a known pathogenic variant in a validated cardiomyopathy-associated gene, indicating that a large proportion of patients have a gene-elusive cause of ACM.

Myoferlin (MYOF) is a type II transmembrane protein with multiple C2 domains, which is highly expressed in skeletal and cardiac muscle (*3*). Prior work has shown that loss of MYOF results in compromised myotubule fusion, smaller muscles, T-tubular disorganization, and inadequate response to injury compensated by enhanced fibrofatty replacement (*4*, *5*).

Mechanistically, MYOF facilitates receptor recycling by promoting receptor internalization, formation of recycling endosomes, and return of receptors to the cell membrane. In the absence of MYOF, endosomes instead fuse with lysosomes, leading to degradation of their contents, including extracellular receptors, rather than recycling them (*5*). MYOF also participates in excitation-contraction coupling in skeletal muscle (*6*, *7*). Recent case reports suggest that *MYOF* variants (such as p.D1575H) may be associated with cardiomyopathy and arrhythmogenesis (*8*), specifically ARVC (*9*). Despite significant insights into the role of MYOF in skeletal muscle, the molecular mechanisms of MYOF disruption have not been studied in the context of cardiomyopathy and arrhythmogenesis. Specifically, evidence supporting *MYOF* as a causative gene in ACM remains limited.

Here, we describe a heterozygous missense variant in *MYOF* (c.4960G>A; p.G1654S) in a family with highly variable expression of ARVC/ALVC and heart failure with reduced ejection fraction (HFrEF). We also identified three additional unrelated cases of cardiomyopathy and arrhythmia in MYOF p.G1654S carriers. We showed that MYOF p.G1654S protein expression is reduced via accelerated lysosomal degradation. To assess the mechanism of MYOF loss in the heart, we generated patient-specific inducible pluripotent stem-derived cardiomyocyte lines (iPSC-CMs) and characterized pro-arrhythmogenic phenotypes via calcium (Ca^2+^) mishandling. Our findings define a direct interaction between MYOF and the L-type voltage-gated Ca^2+^ channel CaV1.2, disruption of which reduces L-type Ca^2+^ current. Verapamil can therapeutically stabilize this disruption of the MYOF-CaV1.2 interaction in p.G1654S carriers. We also established a cardiac-specific *Myof* ^+/-^ model, which revealed cardiomyopathy phenotypes and electrical disruptions. In summary, we define a non-desmosomal mechanism of ACM. In doing so, this work could provide a basis for reclassifying gene-elusive ACM and identifying MYOF as a viable therapeutic target.

## Results

### Variants in *MYOF* are associated with ACM

We identified a family with a variable presentation of arrhythmogenic cardiomyopathy (**Fig. 1A, Fig. S1**) who underwent clinical genetic testing that failed to reveal any pathogenic variants in established ACM genes. The two affected siblings have differential ventricular involvement and arrhythmia burden: one fulfilling criteria for ARVC (II.2), and the other for ALVC (II.1), notably also presenting with heart failure with reduced ejection fraction (HFrEF) (**Fig. 1B**). Their parent (I.1) suffered from non-ischemic cardiomyopathy (CM) with ventricular arrhythmias and a family history of sudden death. To identify novel causative variants associated with disease, we performed whole-exome sequencing (WES) and specifically identified variants that segregated with affected family members. We then applied thresholds to filter candidate variants specific to rare cardiomyopathy diagnosis, such as mean allele frequency (MAF) adjusted for rare disease and REVEL, a missense variant pathogenicity prediction algorithm (MAF<10^-4^, REVEL>0.6) (**Fig. 1C**). Of the three final candidate variants, two, *MYOF* and *FAT1,* were expressed in cardiac tissue. Only *MYOF* had strong evidence linking it to arrhythmogenesis and cardiomyopathy (*8*, *9*) as well as mechanistic evidence for a role in striated muscle (*3*, *4*, *6*) and excitation-contraction coupling (*7*). MYOF p.G1654S was confirmed via Sanger sequencing (**Fig. 1D**) and was highly conserved in vertebrate species (**Fig. 1E**). Additionally, we mined highly phenotyped human biobanks for evidence of *MYOF* contribution to cardiac disease (*10*, *11*). We identified several patients with MYOF p.G1654S: One in the University of Pennsylvania Biobank (Penn Biobank) (*10*) and five additional unrelated patients in Vanderbilt Biobank (BioVU) (*11*) (**Table S1**). We confirmed the absence of any ClinVar 2* P/LP variants (*12*) in 169 genes associated with cardiomyopathy and arrhythmia. Of these patients, four have had a significant cardiomyopathy history, including an LV assist device (LVAD) and subsequent transplant, a history of nonischemic cardiomyopathy with ventricular tachycardia (VT), a resolved history of stress cardiomyopathy, and LV hypertrophy without coronary artery disease (CAD). The other two patients carrying the variant did not undergo echocardiograms or electrocardiograms (ECGs). These findings support the notion that MYOF p.G1654S is strongly associated with cardiomyopathy and arrhythmias in the human population.

**Figure 1.**
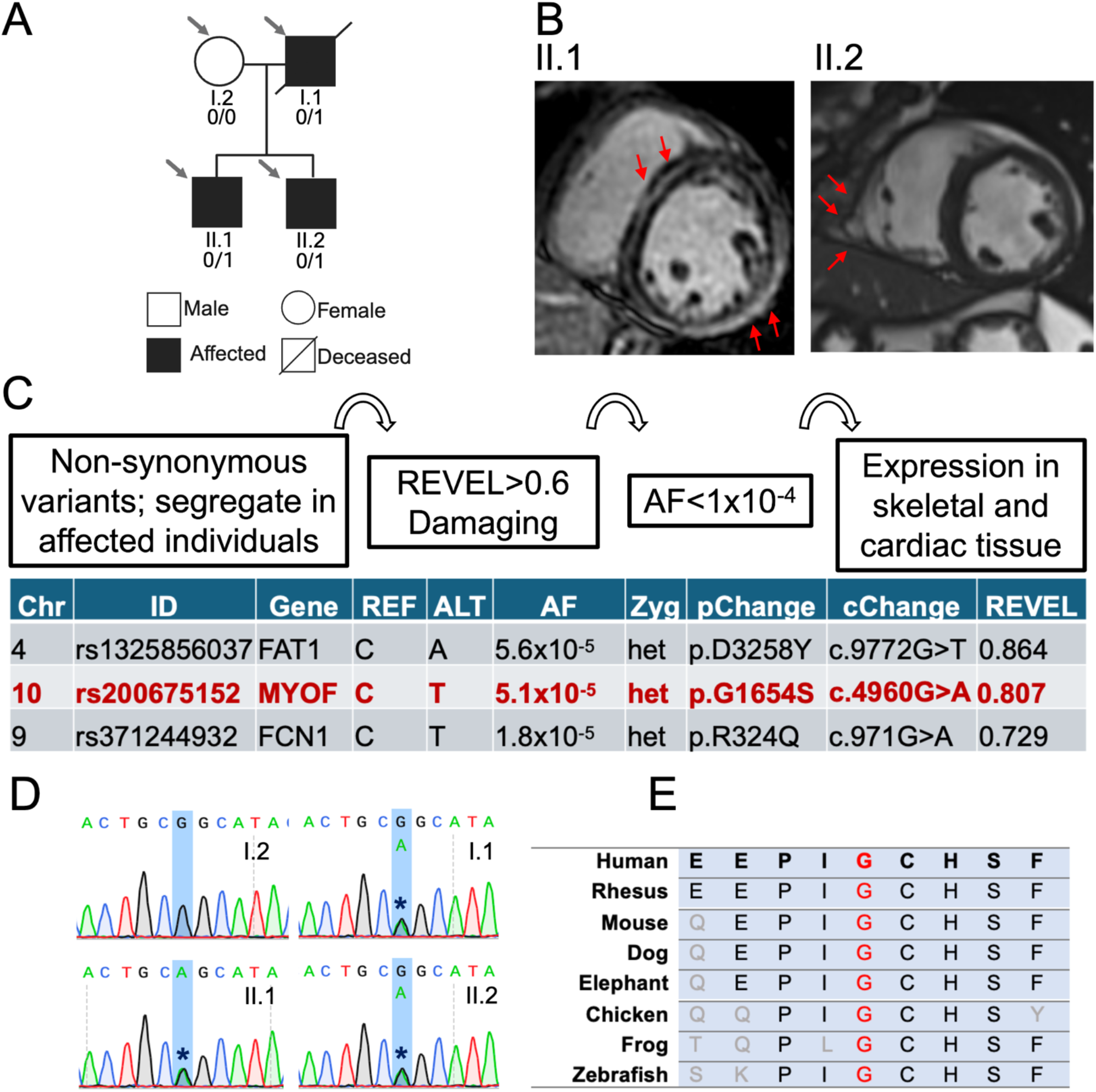
Family affected by ARVC and ALVC. **A.** Arrows point to family members who were investigated. Black squares, affected by cardiac disease; half-filled in black squares, cases of sudden death; crossed out, deceased; 0/0 p.G1654S negative, 0/1 heterozygous for p.G1654S; square, male; circle, female. Pedigree was abridged for patient privacy. Full pedigree is available upon request. **B.** Representative cardiac magnetic resonance imaging (CMR) from II.1 and II.2. II.1 is an inversion recovery sequence, and II.2 is a steady-state free precession sequence. The Red arrow points to a ring-like LGE in the LV of II.1 and the RV aneurysm in the RV of II.2. **C.** Variants were identified based on their segregation with the affected family members (I.1, II.1, II.2). Candidate missense variants were filtered based on REVEL (rare exome variant ensemble learner) prediction (REVEL>0.6) (*77*) and mean allele frequency (MAF) adjusted for rare disease (AF<10^-4^) (*78*). Variants in *MYOF* and *FAT1* were prioritized based on expression in cardiac tissue. There have been no reports linking FAT1 to arrhythmogenic cardiomyopathy or related cardiac phenotypes. Chr (chromosome), ID (identification number of the variant), REF (reference allele), ALT (alternative allele), AF (allele frequency), pChange (protein change), cChange (chromosome change). **D.** Sanger sequencing to confirm MYOF c.4960G>A. **E.** Conservation analysis. Glycine 1654 in red is conserved in all species.

### MYOF p.G1654S undergoes lysosomal degradation and depletes wild-type MYOF

To investigate the mechanisms of cardiomyopathy *in-vitro*, we established inducible pluripotent stem cell lines (iPSC) from each affected individual (I.1, II.1, II.2) and an unaffected control (**Fig. 2A, Fig. S2**). iPSCs were differentiated into ventricular-like cardiomyocytes and showed characteristic sarcomere alignment (**Fig. 2B**). To investigate the effects of *MYOF* variants, we performed molecular dynamics (MD) simulations to assess the stability of the C2F domain in the wild type (WT), p.G1654S, and p.D1575H (*9*) states. Three independent MD simulations were conducted to quantify structural displacement relative to the reference structure. Modeling of the WT C2F domain revealed a stable conformation (RMSD ≈ 3 Å). In contrast, introduction of either p.G1654S or p.D1575H (*9*) into the WT structure resulted in a marked decrease in protein stability (RMSD=4–6 Å for p.G1654S; RMSD = 3–9 Å for p.D1575H) (**Fig. S3**).

**Figure 2.**
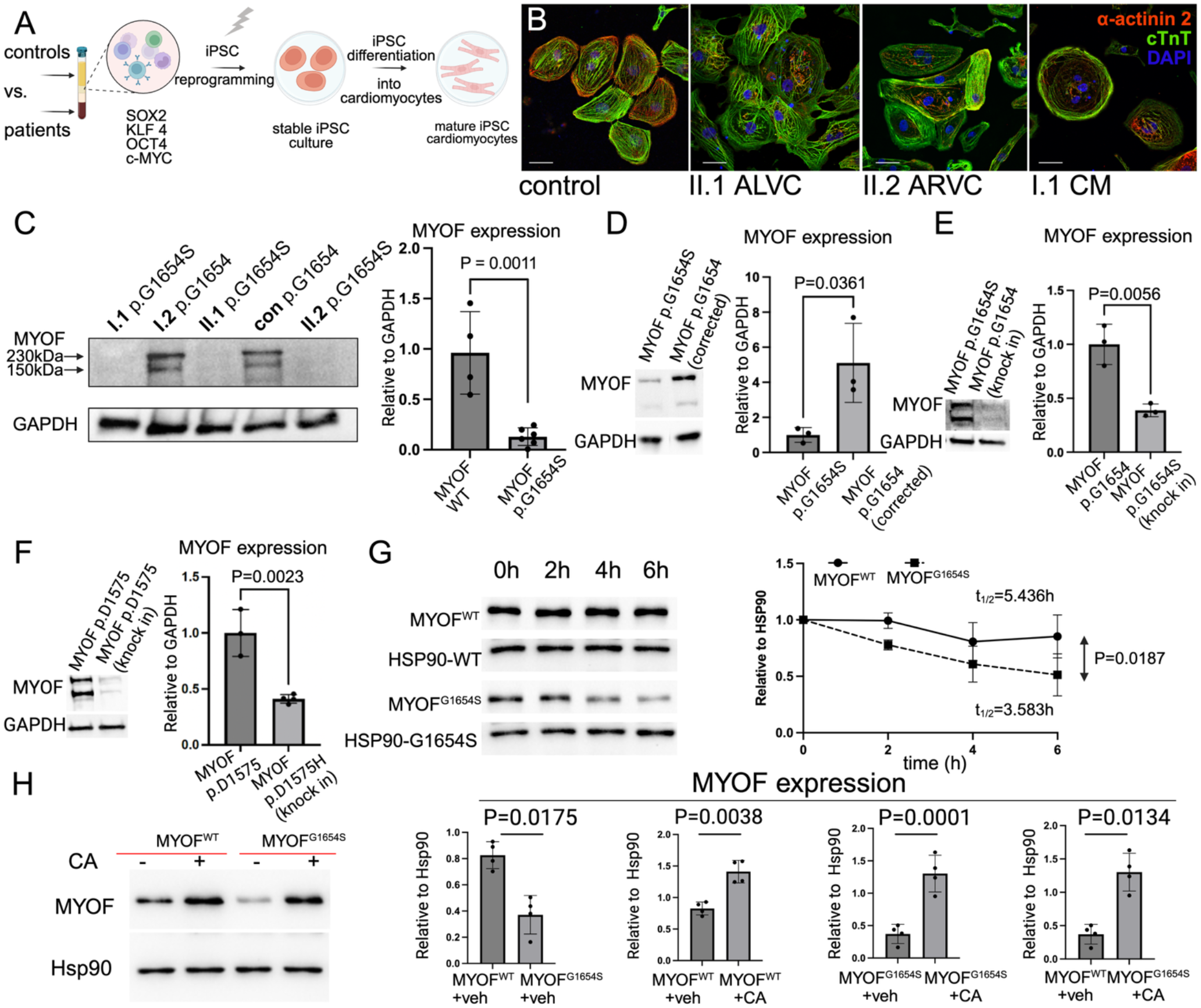
MYOF p.G1654S is degraded faster through a lysosomal pathway. **A.** Samples from I.1, II.1, II.2, and unaffected control were collected and reprogrammed into stable iPSCs. iPSCs were differentiated into cardiomyocytes, which were utilized for experiments. **B.** Immunofluorescent staining of iPSC-CMs from control, ALVC II.1, ARVC II.2 and CM I.1 patients, showing sarcomere structure (red; alpha-actinin2, green; cardiac troponin T; blue; DAPI). **C.** Representative Western blot of MYOF levels in I.1, II.1, II.2 (MYOF p.G1654S carriers) compared to I.2 and other controls (patients n=6 vs. control n=4). **D.** Correction of the MYOF p.G1654S variant in II.1 iPSCs and subsequent differentiation into cardiomyocytes restored MYOF protein expression (corrected n=3 vs. control n=3). Approximately a fivefold increase in MYOF protein levels compared to uncorrected patient cells. **E.** Introduction (knock-in) of the p.G1654S variant into control iPSCs, followed by cardiomyocyte differentiation. MYOF expression decreased by more than twofold (knock-in n=3 vs. control n=3). **F**. Knock-in of p.D1575H reduced MYOF protein levels by more than twofold (knock-in n=3 vs. control n=3). All protein levels were normalized to the GAPDH loading control. Experiments were performed using three independent iPSC-CM differentiation samples. A two-tailed unpaired Student’s *t*-test was used to assess statistical significance. **G.** Cycloheximide (CHX) chase assay. HEK293T cells were transfected with either MYOF^WT^ or MYOF^G1654S^ and treated with 300ug/mL CHX at t=0. Lysates were harvested every 2 hrs for 6 hrs. Representative western blots normalized to the housekeeping protein HSP90 are shown (MYOF^G1654S^ n=5 vs. MYOF^WT^ n=5). MYOF^WT^ ^and^ MYOF^G1654S^ were both normalized to 1 at t=0. Half-lives were determined by fitting a one-phase decay model. Slope difference was tested with a two-way repeated-measures ANOVA (p=0.0187). **H.** Lysosomal inhibition rescue assay. HEK293T cells were transfected with either MYOF^WT^ or MYOF^G1654S^ and treated with either 100nM Concanamycin A (CA) or DMSO vehicle for 24 hrs. n=4 samples were used in each condition. P values were calculated by two-way ANOVA followed by Tukey’s multiple comparison test.

To determine whether MYOF protein expression is altered in carriers of the p.G1654S variant, we compared iPSC-CMs from non-carrier controls to all affected carriers (I.1, II.1, II.2). Quantitative analysis of the major MYOF isoform (230kDa) showed a greater than 50% decrease in protein expression in patient samples compared to controls (**Fig. 2C**). Isogenic CRISPR/Cas9 correction of p.G1654S in II.1 ALVC iPSC-CMs restored MYOF protein expression (**Fig. 2D**). Introduction of p.G1654S into control iPSC-CMs similarly reduced MYOF protein levels (**Fig. 2E**), confirming that the p.G1654S variant is both necessary and sufficient to disrupt MYOF expression. Given that the p.D1575H variant (*9*) resides in the same functional C2F domain as p.G1654S, we hypothesized that it might similarly reduce MYOF protein expression through a similar mechanism. Indeed, introduction of p.D1575H (*9*) into control iPSC-CM resulted in MYOF depletion (**Fig. 2F**).

To investigate whether the reduction in protein expression is due to enhanced degradation of MYOF p.G1654S, we performed a cycloheximide (CHX) chase assay. HEK293T cells were transfected with either MYOF^WT^ or MYOF^G1654S^ expression constructs. After 6 hours of CHX treatment, MYOF^G1654S^ had a shorter half-life (t_1/2_=3.583) than MYOF^WT^ (t_1/2_=5.436). MYOF^G1654S^ was also degraded more rapidly than MYOF^WT^ (**Fig. 2G**).

MYOF has been extensively shown to localize to late endosomes and lysosomes (*13–15*); therefore, we hypothesized that lysosomal degradation contributes to accelerated MYOF^G1654S^ turnover. We treated MYOF^WT^ or MYOF^G1654S^ with Concanamycin A (CA), an inhibitor of vacuolar-type H+-ATPase (*16*) and lysosomal degradation. At baseline, steady-state MYOF^G1654S^ levels were lower than MYOF^WT^. Treatment with CA upregulated MYOF^WT^ and MYOF^G1654S^ expression compared with vehicle-only treatment. Upregulation of MYOF^G1654S^ after CA treatment brought the MYOF levels up to levels comparable to those of MYOF^WT^ (**Fig. 2H**). These results show that MYOF^G1654S^ is less stable and degrades faster via a lysosome-specific mechanism, and that lysosomal degradation inhibitors can rescue MYOF^G1654S^ downregulation.

The greater-than-50% reduction of MYOF protein observed in heterozygous p.G1654S carrier iPSC-CMs (**Fig. 2C**) exceeded the predicted ∼50% loss by depletion of the variant allele product alone, raising the possibility that MYOF^G1654S^ additionally destabilizes MYOF^WT^ in *trans*. To test this directly, we co-expressed a fixed amount of MYOF^WT–FLAG^ (500 ng) together with increasing amounts of MYOF^G1654S–HA^ (0–2000 ng) in HEK293T cells. MYOF^G1654S–HA^ accumulated in a dose-dependent manner (**Fig. S4C**), and this was accompanied by progressive, dose-dependent depletion of MYOF^WT–FLAG^ in the same lysates (**Fig. S4D**). This effect was specific to the variant protein; when an identical titration was performed with MYOF^WT–HA^ (**Fig. S4E**), MYOF^WT–FLAG^ levels remained unchanged across the entire dose range (**Fig. S4F**), excluding increasing plasmid load or competition for the transcriptional and translational machinery as an explanation. Conversely, titrating MYOF^WT–FLAG^ (0–2000 ng) against a fixed amount of MYOF^G1654S–HA^ (125 ng) did not restore MYOF^G1654S^ protein levels (**Fig. S4G–H**), indicating that the destabilizing effect is unidirectional, where MYOF^G1654S^ depletes MYOF^WT^, while excess MYOF^WT^ does not rescue the variant.

To determine whether this *trans* effect reflects a physical association between MYOF molecules, we co-expressed differentially tagged MYOF constructs and performed anti-FLAG immunoprecipitation followed by anti-HA immunoblotting. MYOF co-precipitated with itself in all pairwise combinations tested, and MYOF^G1654S^ retained the capacity to associate with MYOF^WT^in both tag orientations (**Fig. S4I**), providing a physical basis through which the destabilized variant can engage and co-deplete MYOF^WT^. Together, these results provide direct biochemical evidence that MYOF p.G1654S acts in a dominant-negative fashion on the wild-type allele product, and account for the greater-than-haploinsufficient loss of MYOF observed in heterozygous carriers.

### MYOF mediates arrhythmogenesis and calcium cycling

To elucidate the pathways reprogrammed in primary patient iPSC-CMs carrying the MYOF p.G1654S (I.1, II.1, II.2), we performed RNA sequencing on three independent samples and compared them to the control (**Fig. 3A**). To better understand transcriptome reprogramming, we performed Gene Ontology (GO) enrichment analysis (*17*), which revealed top terms related to remodeling of extracellular space (**Fig. 3B**), a well-established mechanism of ACM (*18–21*). Further examination of GO terms identified at least 16 terms specific to Ca^2+^ signaling (**Fig. 3C**), prompting us to investigate more refined molecular function (MF) GO terms. MF terms were linked to ion cycling and channel activity, including ‘monoatomic channel activity’, ‘metal ion transmembrane transporter activity’, and ‘monoatomic cation channel activity’. We utilized ClusterProfiler to generate a network of biological terms (*22*), which revealed a prominent cluster enriched for voltage-gated ion channels and Ca^2+^ handling, suggesting that Ca^2+^ homeostasis may be disrupted in iPSC-CMs from MYOF p.G1654S carriers (**Fig. 3D**).

**Figure 3.**
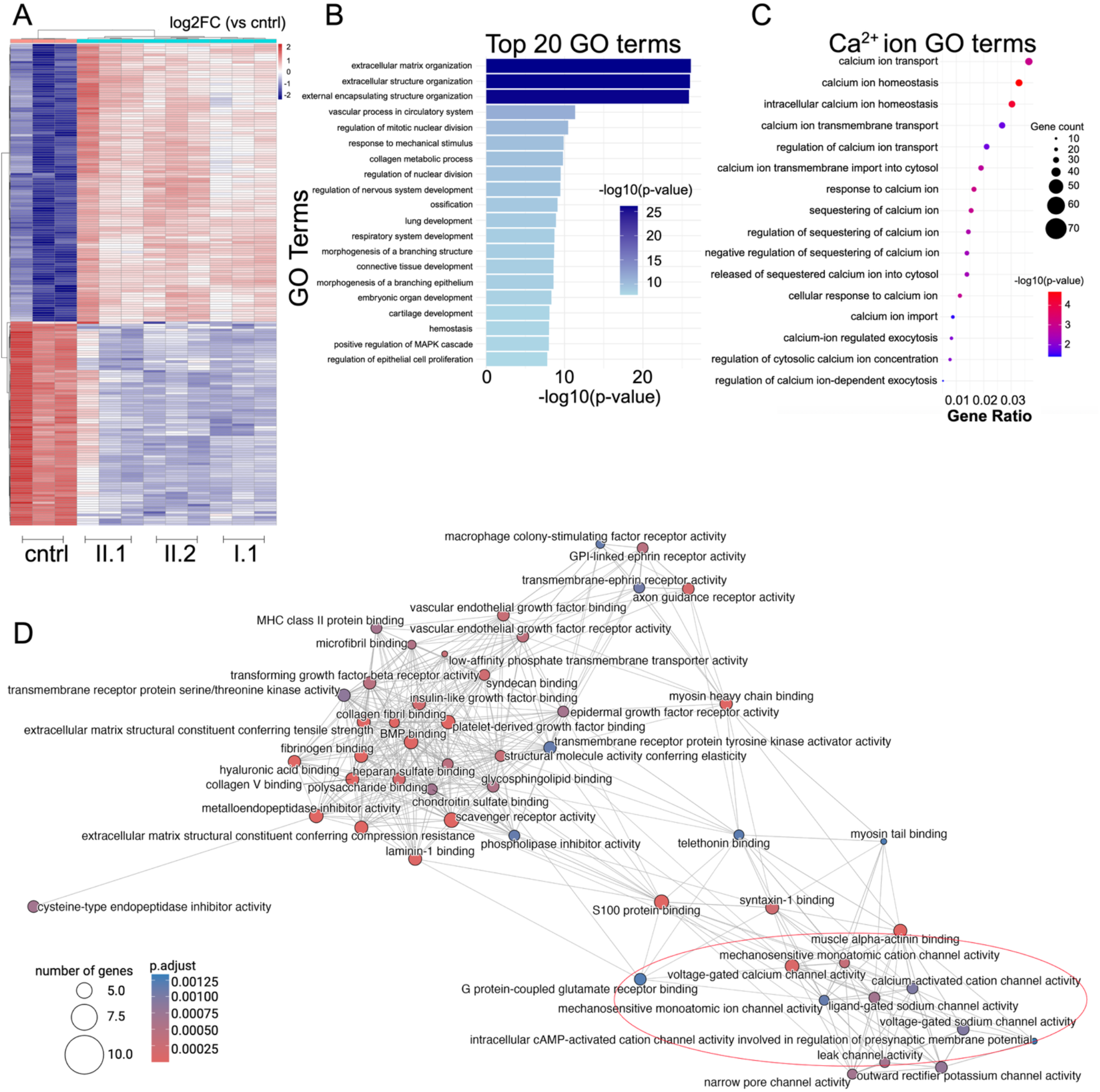
RNA sequencing analysis of patient-specific iPSC-CMs reveals top pathways related to calcium signaling. **A.** All differentially expressed genes (DEGs) in iPSC-CMs from ALVC II.1, ARVC II.2, and CM I.1 compared to control (cntrl). Only DEGs that were consistently reversed in affected individuals (up- or downregulated) compared to controls. Three samples from each patient were independently sequenced. Colors represent log2FC: blue, downregulated DEGs; red, upregulated DEGs. Only DEGs meeting adjusted p<0.05 are shown. **B.** Gene ontology (GO) analysis of all DEGs. Only the top 10 terms are shown. Bars are color-coded based on -log10(p-value). **C.** Calcium-specific terms selected from the GO analysis. GO terms plotted as a function of descending gene ratios. The relative size of the dots represents gene count. Dots are color-coded based on -log10(p-value). **D.** Molecular function (MF) GO terms analysis plotted as a network using ClusterProfiler (*22*). Terms are grouped by relative biological similarity and function. All MF terms are shown. Terms related to ion handling are circled in red. The size of dots reflects the number of genes in each GO term. Dots are color-coded based on the adjusted p-value. Lines show similarity between GO terms based on the Jaccard index.

These findings, along with the fact that MYOF is a C2-domain-containing, Ca^2+^-binding protein known to regulate Ca^2+^ cycling (*7*), supported the notion that the MYOF p.G1654S could contribute to Ca^2+^ mishandling and underlie the arrhythmia observed in affected family members. To investigate, we utilized patient-specific iPSC-CMs (I.1, II.1, II.2) and compared their Ca^2+^ handling to that of unaffected controls. Our analyses revealed that MYOF p.G1654S iPSC-CMs exhibited significantly reduced Ca^2+^ amplitude, elevated diastolic baseline Ca^2+^ levels, and prolonged Ca^2+^ transient duration (CaT) at 50% and 90% (**Fig. 4A-B**). These results are consistent with the established model of Ca^2+^ overload, prolonged CaT, and heart failure (*23–25*), indicating that MYOF p.G1654S disrupts one or more key Ca^2+^ handling mechanisms.

**Figure 4.**
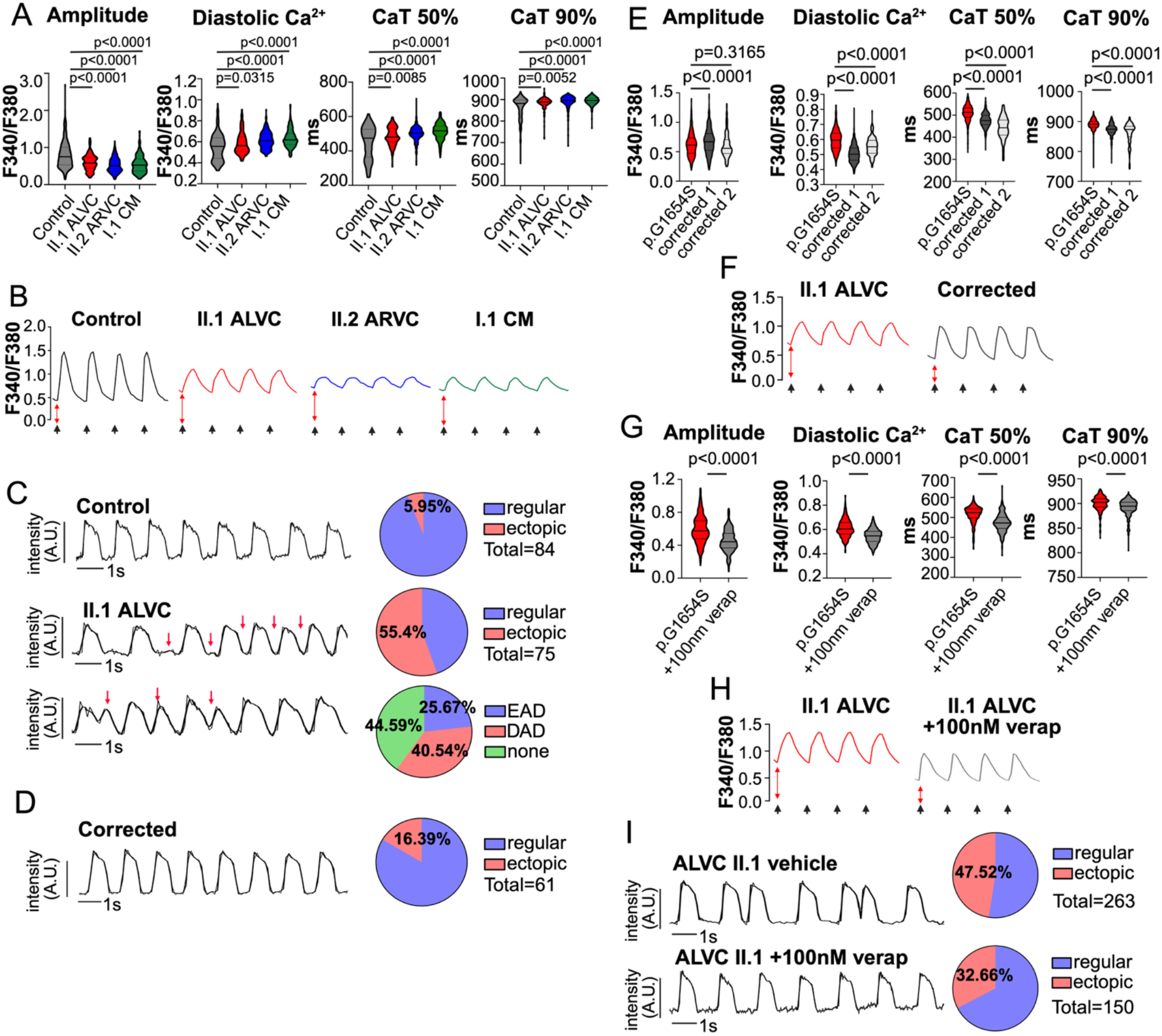
**MYOF p.G1654S controls Ca**^2+^ **handling and arrhythmogenesis. A.** Calcium mishandling in iPSC-CM II.1, II.2, I.1 compared to control after 30 days of differentiation. Decreased amplitude, elevated diastolic Ca^2+,^ and prolonged Ca^2+^ transient duration (CaT) at 50% and 90% in patient iPSC-CM (ALVC II.1, ARVC II.2, CM I.1) compared to control. For each condition, control n=304; ALVC II.1 n=219, ARVC II.2 n=231, CM I.1 n=259 cells were used. **B.** Representative traces showing control, ALVC II.1, ARVC II.2, CM I.1 Ca^2+^ dynamics. Tick marks indicate 1 Hz stimulation pacing. **C.** Arrhythmia burden in ALVC II.1 iPSC-CM compared to control via optical action potential (AP) traces. Ectopic beats were defined as EAD or DAD. Arrhythmia burden is shown as a fraction of cells with ectopic beats (5.95%). At least n=84 was utilized for control analysis. Red arrows represent aberrant beats (EAD or DAD). Of the 55.4% ALVC II.1 iPSC-CMs with ectopic rhythm, abnormalities were classified into EAD (25.67%), DAD (40.54%), or none (44.59%). Some cells showed both DADs and EADs, resulting in a total >100%. At least n=75 cells were utilized for the analysis of ALVC II.1. Cells were not paced to unmask the intrinsic arrhythmia phenotype. **D.** AP traces showing isogenic CRISPR/Cas9 correction of MYOF p.G1654S in ALVC II.1 iPSC-CM. The fraction of cells with ectopic beats is represented as a percent of the whole (16.39%). At least n=61 cells were utilized for the analysis. Cells were not paced to unmask any residual intrinsic arrhythmia phenotype. **E.** Calcium handling dynamics in isogenic CRISPR/Cas9 corrected iPSC-CM II.1. Increased amplitude, decreased diastolic Ca^2+^, and shortened Ca^2+^ transient duration (CaT) at 50% and 90% in patient iPSC-CM. iPSC-CM ALVC II.1 (n=497), isogenic iPSC-CM II.1 p.G1654S corrected clone 1 (n=577), clone 2 (n=168). **F.** Representative traces showing ALVC II.1 and isogenic corrected iPSC-CM. **G.** Ca2+ mishandling rescued with verapamil treatment. Decreased amplitude, decreased diastolic Ca^2+^, and shortened Ca^2+^ transient duration (CaT) at 50% and 90% in patient iPSC-CM treated with verapamil. iPSC-CM ALVC II.1 treated with 100nM verapamil (n=366) compared to vehicle (n=419). **H.** Representative Ca^2+^ traces of verapamil compared to vehicle-treated iPSC-CM ALVC II.1. **I.** Verapamil (verap) rescues arrhythmias in iPSC-CM ALVC II.1. Verapamil reduced arrhythmogenic activity from 47.52% (ALVC II.1 vehicle, n=263) to 32.66% (ALVC II.1 + 100 nM verap, n=150). Cells were not paced to unmask any residual intrinsic arrhythmia phenotype. Violin plots were reported as median ± IQR. Statistical testing was performed under the assumption that each cell was an independent unit of observation. P values were calculated by Kruskal-Wallis test followed by Dunn’s multiple comparison test (**A** and **E**). Mann-Whitney test was used to assess statistical significance in binary comparisons (**G**).

To assess whether ventricular-like iPSC-CMs derived from ALVC II.1 patient exhibited a greater propensity for arrhythmic events, as sequelae of elevated diastolic Ca^2+^ and prolonged CaT, we utilized optical action potential (AP) recordings. Compared to control, ALVC II.1 iPSC-CMs displayed a profound enrichment of early (EAD) and delayed afterdepolarizations (DAD) (ALVC II.1 = 55.4% vs. control = 5.95%) (**Fig. 4C**).

Isogenic CRISPR/Cas9 correction of the MYOF p.G1654S variant resulted in reversal of Ca^2+^ handling defects, including significantly reduced diastolic Ca^2+^, shortened CaT at 50% and 90% (**Fig. 4E-F**). Calcium amplitude was rescued only in one isogenic-corrected clone. Additionally, correction of the MYOF p.G1654S led to a major reduction in arrhythmic activity (ALVC II.1 = 55.4% vs. corrected = 16.39%) (**Fig. 4D**). Collectively, these results further support the conclusion that arrhythmogenesis in MYOF p.G1654S iPSC-CMs is primarily driven by Ca^2+^ overload and prolonged CaT, which together increase the susceptibility to DADs and EADs.

To complement our genetic rescue, we sought to identify a pharmacological agent capable of producing a similar therapeutic effect. We tested several anti-arrhythmic agents targeting Ca^2+^ ion channels. Verapamil, an L-type Ca^2+^ channel blocker with established anti-arrhythmic properties, significantly shortened CaT at both 50% and 90% recovery and lowered diastolic Ca^2+^ levels in iPSC-CMs from II.1 ALVC (**Fig. 4G-H**). Arrhythmogenicity was also rescued with verapamil treatment (**Fig. 4I**). In contrast, treatment with metoprolol or dantrolene did not produce similar Ca^2+^ handling effects (**Fig. S5**).

iPSC-CMs are an established model for patient-specific disease, including ACM (*26*); however, batch-to-batch variability and inconsistent differentiation have previously affected the interpretation of results (*27*). Therefore, to confirm that the iPSC-CM results are reproducible in an independent cardiac cell system, we isolated ventricular cardiomyocytes (NRVM) from day 0-3 neonatal rats and performed silencer RNA knockdown of *MYOF* (si*MYOF* vs. si*NC*). Silencer RNA knockdown efficiency reached 70% (**Fig. 5A**), which was consistent with decreased MYOF protein expression in iPSC-CM harboring MYOF p.G1654S. Calcium traces from si*MYOF* NRVMs showed an increased burden of atypical Ca^2+^ transients despite electrical pacing at 1 Hz (**Fig. 5B**). Optical recording of Ca^2+^ handling in si*MYOF* NRVMs phenocopied our results in iPSC-CM MYOF p.G1654S (**Fig. 5C**). Overall, these results support that MYOF exclusively controls intracellular diastolic Ca^2+^ overload and the duration of CaT, which may precipitate abnormal Ca^2+^ transients and, therefore, intrinsic arrhythmogenic activity.

**Figure 5.**
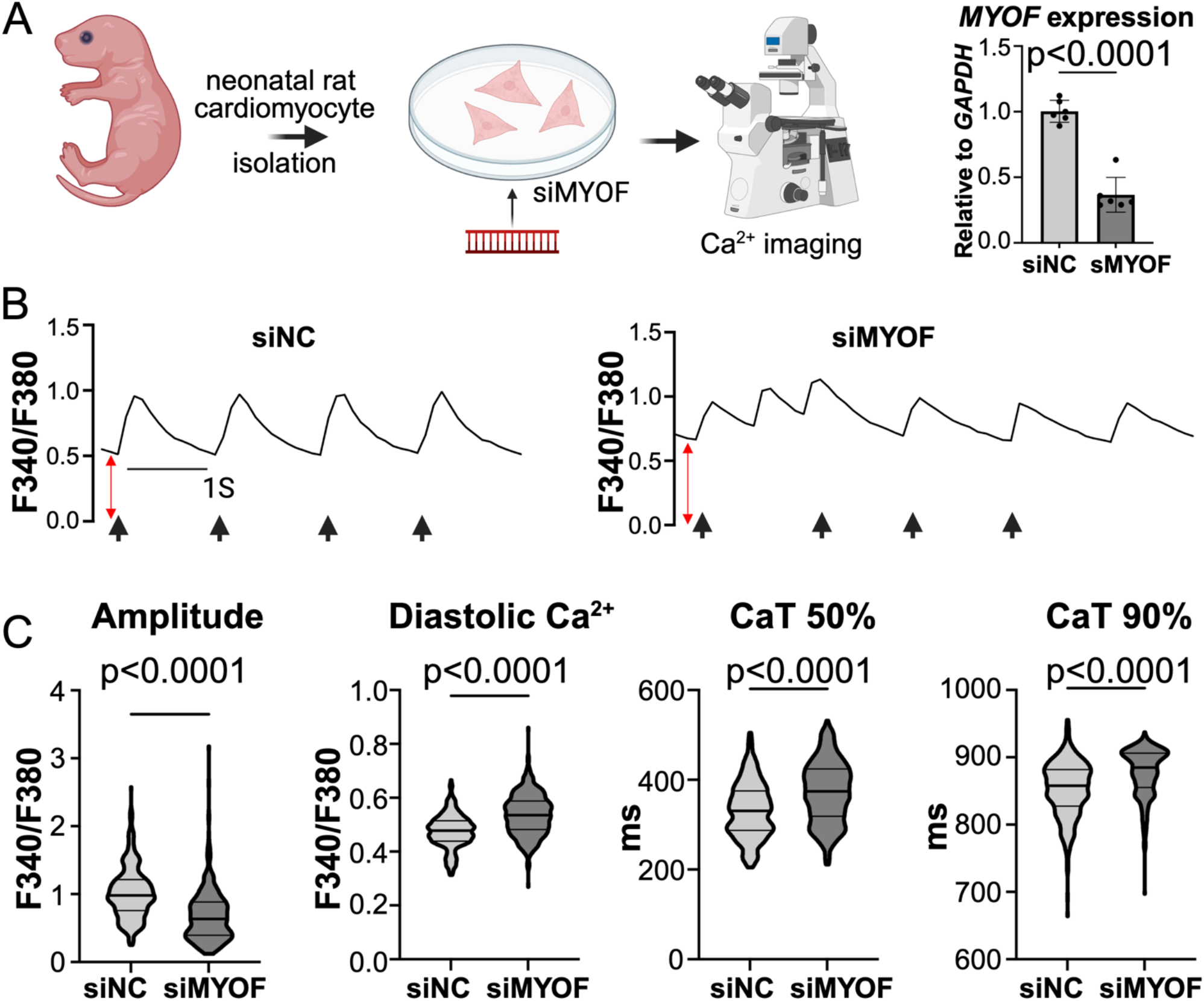
Calcium handling is altered in neonatal rat cardiomyocytes (NRVM) with *MYOF* knockdown. **A.** NRVM were isolated and treated with siRNA specific to *MYOF* (*siMYOF*) or scrambled transcripts (*siNC*) prior to ratiometric Ca^2+^ recording acquisition using Fura-2 AM. Validation of *MYOF* transcript knockdown (*siMYOF* n=6 vs. *siNC* n=6). Knockdown efficiency of *MYOF* averaged 70%. **B.** Representative traces of Ca^2+^ oscillations in *siMYOF* vs. *siNC* NRVMs. Tick marks represent stimulation pacing at 1 Hz. Red arrows show diastolic Ca^2+^ increase in *siMYOF* compared to *siNC*. **C.** The amplitude of Ca^2+^ oscillations was significantly decreased; diastolic Ca^2+^ was significantly elevated, and CaT 50% and 90% were significantly prolonged (*siMYOF* n=382 vs. *siNC* n=408) and were used. Violin plots were reported as median ± IQR. Statistical testing was performed under the assumption that each cell was an independent unit of observation. Mann-Whitney test was used to assess statistical significance.

### MYOF interacts with L-type voltage-gated calcium channel CaV1.2

Previous reports have shown that MYOF plays a role in T-tubule organization in skeletal muscle (*5*). Given the observed CaT phenotype, we hypothesized that MYOF could co-localize with a membrane ion channel mediating Ca^2+^ entry into the cell. Because verapamil rescued Ca^2+^ mishandling and is specific to L-type Ca^2+^ channels, we further investigated the L-type Ca^2+^ channel CaV1.2 as a potential interacting partner of MYOF (*28*). To determine whether MYOF exhibits T-tubule distribution in cardiac tissue and cardiomyocytes, we performed co-immunofluorescence (IF) staining of MYOF with CaV1.2, a T-tubule and sarcolemma marker. In cardiac tissue and ventricular cardiomyocytes, MYOF showed strong co-localization with CaV1.2 (**Fig. 6A-C**). To further investigate this co-localization, we utilized *in silico* modeling with AlphaFold (AF) to align MYOF with CaV1.2 (**Fig. 7A**). Both p.G1654S and p.D1575H are localized to the C2F domain. AF predicted an unusual interaction feature, where a flexible linker encircles the C2F domain, creating a lever-like ring, which we called the ‘linker domain’ (**Fig. S6**). We identified the ‘KFYA’ motif within the MYOF ‘linker domain’, near the C2F domain, that perfectly overlaps with the IQ domain of CaV1.2 (**Fig. 7B-C**). We hypothesized that the linker domain acts as a hinge that an interacting molecule could displace, enabling the CaV1.2 IQ domain to interact with the MYOF C2F domain pocket (**Fig. 7D-G**). In the native state, the MYOF linker domain interacted with its native C2F pocket (**Fig. 7D**); the p.G1654S variant made this interaction less stable by disrupting hydrogen (H) bonds (**Fig. 7E**). In the CaV1.2 ‘bound’ state, the MYOF linker was displaced, allowing for the CaV1.2 to interact with the MYOF C2F (**Fig. 7F**). Introduction of the p.G1654S variant weakened H-bonds with CaV1.2 and introduced steric hindrance (**Fig. 7G**).

**Figure 6.**
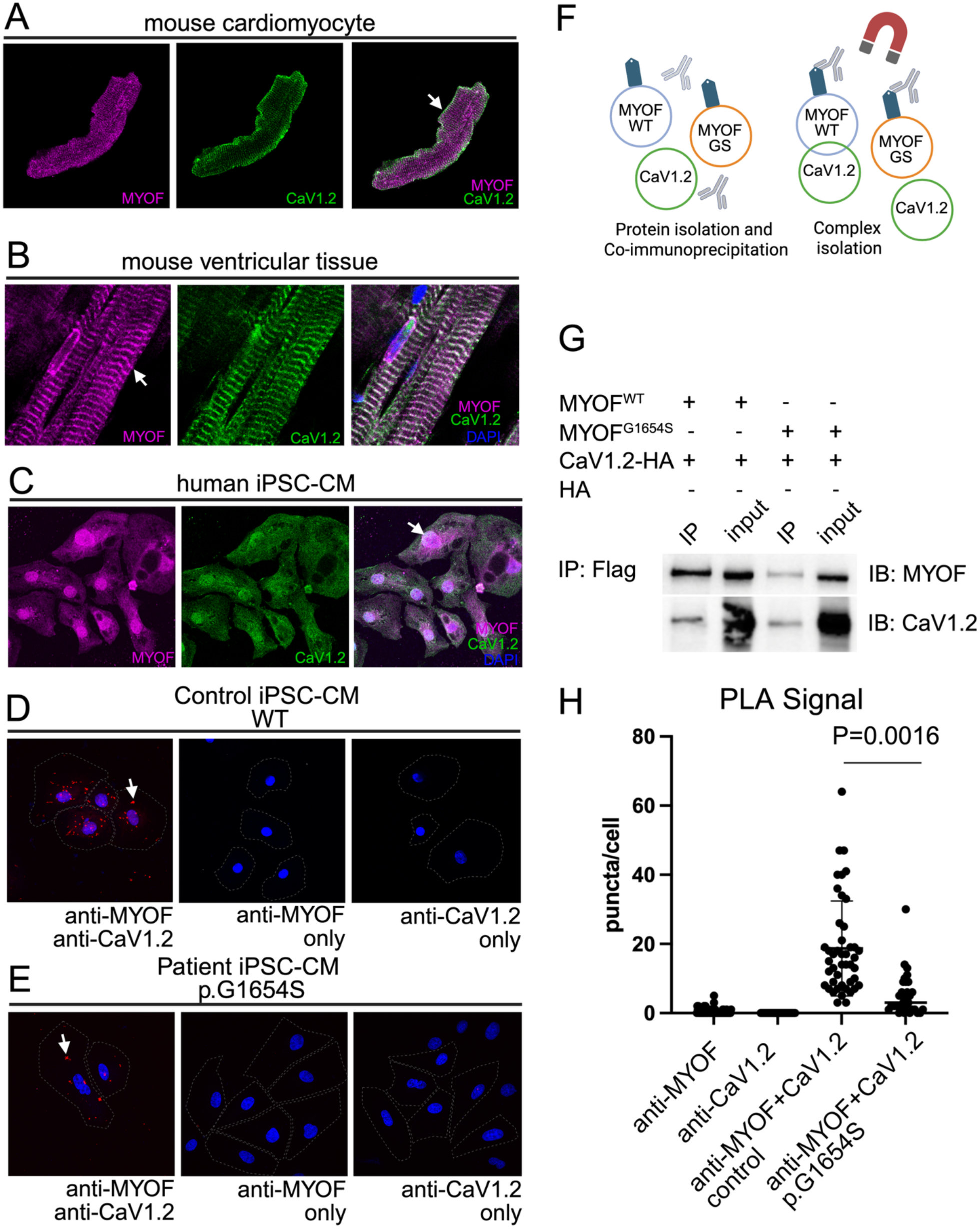
MYOF interacts with CaV1.2. **A.** Dissociated adult mouse cardiomyocytes. MYOF was co-stained with CaV1.2. Overlayed images showing co-localization or lack thereof. MYOF co-localized with CaV1.2. **B.** CaV1.2-MYOF co-localization in isolated adult mouse ventricular tissue. **C.** Human control iPSC-cardiomyocytes. Staining is diffuse due to a lack of mature T-tubules, confirming MYOF co-localization with CaV1.2 rather than with myotubules. Nuclear localization of MYOF and CaV1.2 may represent transcription factor activity. **D and E.** Proximity ligation assay (PLA) showing MYOF-CaV1.2 interaction. PLA was performed in control versus patient iPSC-CM ALVC II.1 to evaluate whether there is a change in the MYOF-CaV1.2 interaction. Compared to control, patient MYOF p.G1654S iPSC-CMs had decreased PLA hybridization signal (control anti-MYOF+anti-CaV1.2, n=43, compared to patient anti-MYOF+anti-CaV1.2, n=31; p=0.0016). Negative controls were established by incubating samples with only anti-MYOF or anti-CaV1.2 antibodies. Negative controls were established as follows: anti-MYOF only, n=74 cells (averaged control and patient samples); anti-CaV1.2 only, n=53 (averaged control and patient samples). The PLA signal was absent in all negative control samples. **F.** Co-immunoprecipitation of MYOF with CaV1.2. MYOF^WT-FLAG^ or MYOF^G1654S-FLAG^ expression plasmids were co-transfected with the full-length α-subunit of CaV1.2 to assess their interaction. MYOF was pulled down via anti-FLAG magnetic beads. **G.** Representative Western blot showing the pull-down assay. MYOF^WT^ co-immunoprecipitated with CaV1.2. MYOF^G1654S^ co-immunoprecipitated less with CaV1.2. **H.** Quantification of PLA signal with ImageJ. P values were calculated by Kruskal-Wallis test followed by Dunn’s multiple comparison test.

**Figure 7.**
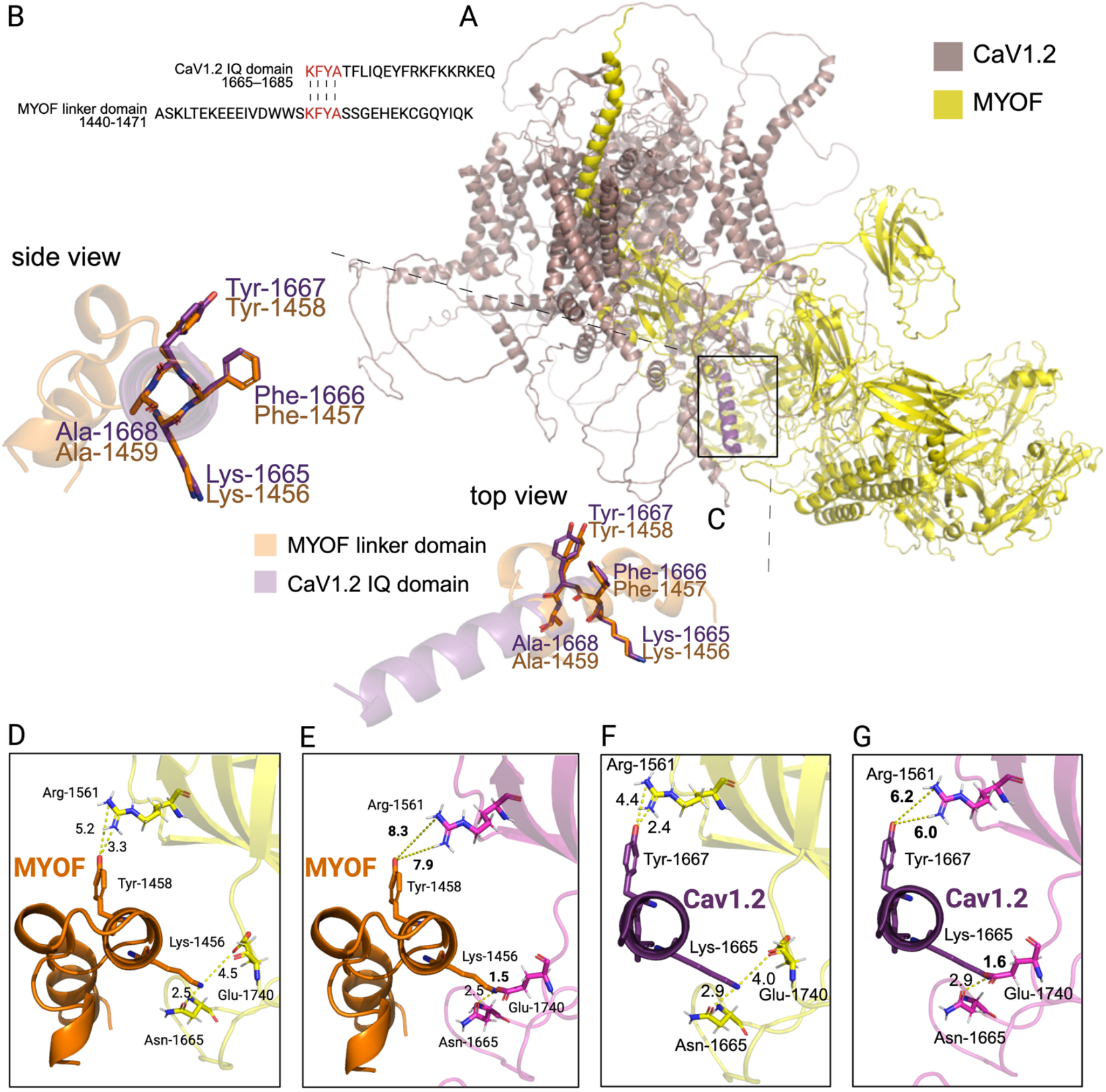
Computational modeling of MYOF-CaV1.2 interaction. **A.** MYOF and CaV1.2 were generated using AlphaFold and aligned in PyMOL. A motif ‘KFYA’ was aligned between the MYOF linker domain (1440-1471) and the CaV1.2 IQ domain (1665-1685). **B**. Modeling of MYOF and CaV1.2 interaction via linker and IQ domains. Cartoon representation of the top and side view alignment between the MYOF linker domain and the CaV1.2 IQ domain. Amino acids that overlap between the two structures are shown as follows: MYOF, Lysine-1456 (Lys), Phenylalanine-1457 (Phe), Tyrosine-1458 (Tyr), Alanine-1459 (Ala), and CaV1.2, Lys-1665, Phe-1666, Tyr-1667, Ala-1668. **D**. PyMOL modeling showing the MYOF linker domain interacting with the C2F WT pocket. Linker domain Tyr-1458 interacts with C2F Arginine-1561 (Arg) (H-bonds, 5.2 Å, 3.3 Å); linker domain Lys-1456 interacts with C2F Asparagine-1665 (Asn) (H-bond, 2.5 Å) and Glutamine-1740 (Glu) (H-bond, 4.5 Å). **E**. PyMOL modeling showing altered interaction between the linker domain and the C2F p.G1654S mutant pocket. H-bond distance is increased between linker domain Tyr-1458 and C2F Arg-1561 (H-bonds, 8.3 Å, 7.9 Å); steric hindrance is acquired between linker domain Lys-1456 and C2F Glu-1740 (H-bond, 1.5 Å). Interaction between Lys-1456 and Asn-1665 is unchanged (H-bond, 2.5 Å). **F**. PyMOL modeling showing a putative interaction between CaV1.2 IQ domain as it displaces MYOF linker domain and interacts with the C2F via Tyr-1667 and Arginine-1561 (Arg) (H-bonds, 4.4 Å, 2.4 Å); Lys-1665 and Asn-1665 (H-bond, 2.9 Å); Glu-1740 (H-bond, 4.0 Å). **G**. PyMOL modeling showing altered interaction between the CaV1.2 IQ domain and the C2F p.G1654S mutant pocket. H-bond distance is increased between CaV1.2 Tyr-1667 and C2F Arg-1561 (H-bonds, 6.0 Å, 6.2 Å); steric hindrance is acquired between CaV1.2 Lys-1665 and C2F Glu-1740 (H-bond, 1.6 Å). Interaction between CaV1.2 Lys-1665 and C2F Asn-1665 is unchanged (H-bond, 2.9 Å).

To validate this interaction *in vitro*, we performed a proximity ligation assay (PLA); MYOF interacted significantly more with CaV1.2 in control iPSC-CMs than in patient II.1 ALVC iPSC-CMs harboring MYOF p.G1654S (**Fig. 6D-E, H**). Additionally, we performed co-immunoprecipitation (Co-IP) analysis by overexpressing MYOF^WT-FLAG^ and MYOF^G1654S-FLAG^ in HEK293T cells with or without the presence of full-length CaV1.2^HA^ (**Fig. 6F**). We detected robust co-immunoprecipitation of CaV1.2 with MYOF^WT^. In contrast, MYOF^G1654S^ interacted less with CaV1.2 (**Fig. 6G**) (see controls in **Fig. S4)**. In conclusion, our *in silico*, PLA, and Co-IP analyses showed that the p.G1654S variant impaired the MYOF–CaV1.2 interaction.

### MYOF loss-of-function promotes arrhythmogenic cardiomyopathy *in vivo*

Systemic *Myof* knockout (*Myof* KO) mice have previously been shown to have smaller skeletal muscles and acquire fatty infiltrate (*4*, *5*). A cardiac-specific *Myof* KO model has not been previously generated and assessed for cardiomyopathy and arrhythmia phenotypes. Supported by clinical and iPSC-CM evidence that patients with pathogenic *MYOF* variants have compromised ventricular function, including reduced LVEF in the ALVC (II.1) proband, we evaluated cardiac function in heterozygous cardiac-specific *Myof* ^+/-^ compared with *Myof* ^+/+^ controls. *Myof* ^+/-^ mice were generated using the *Myh6*-Cre cardiac-specific system and validated per established protocol (**Fig. S7**). Male *Myof* ^+/-^ mice exhibited significant contractile dysfunction beginning at 3 months of age, characterized by markedly reduced left ventricular (LV) fractional shortening (FS) and EF (**Fig. 8A-B, Fig. S8A**). Additionally, both systolic and diastolic diameters were significantly increased, indicating a predominant heart failure (HF) phenotype with LV dilation (**Fig. 8A-B**). In contrast, female *Myof* ^+/-^ mice displayed an attenuated phenotype, with FS showing a trend toward reduction and a trend toward increase in systolic and diastolic diameters (**Fig. S8B**).

**Figure 8.**
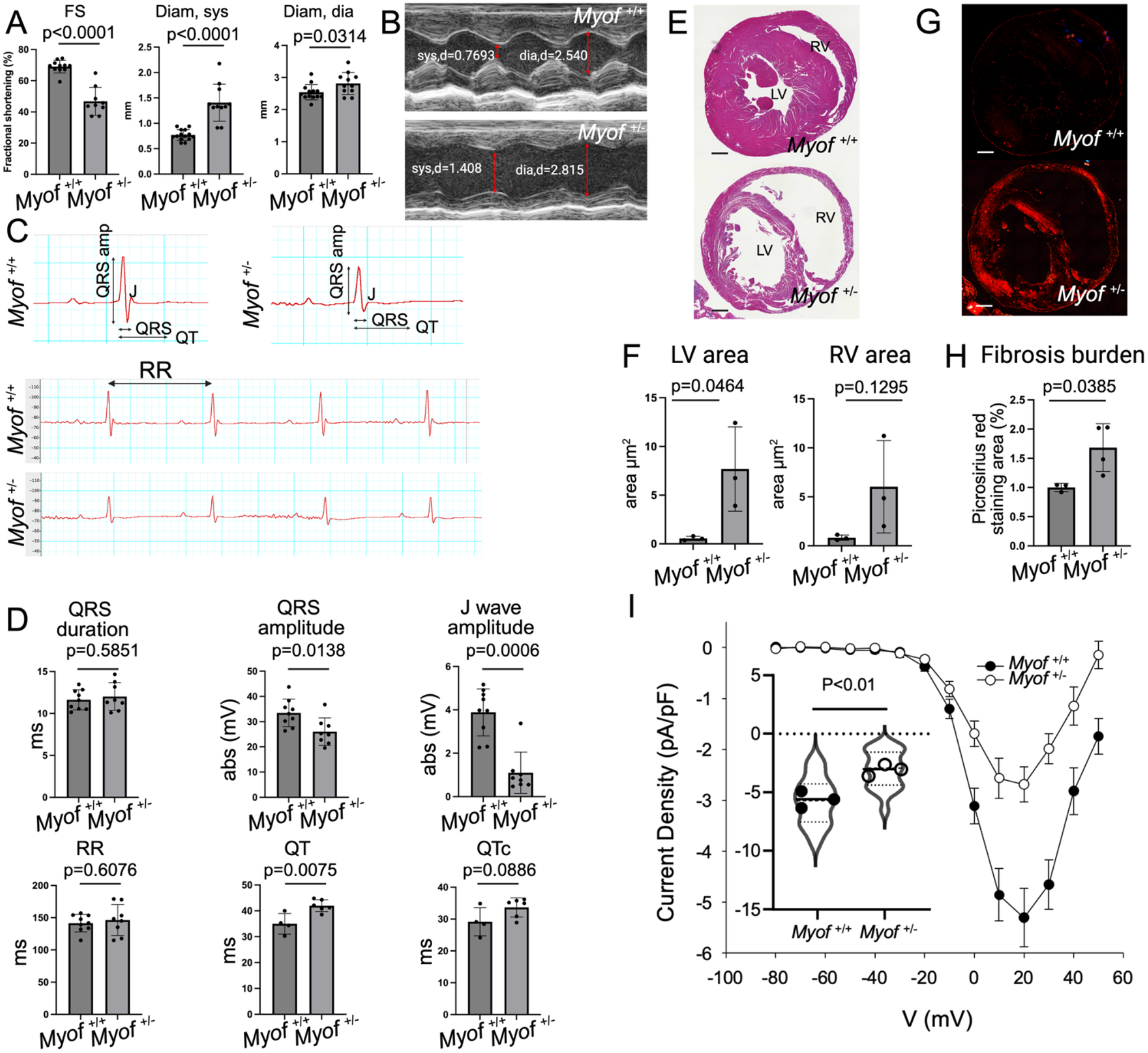
Contractile and electrical function in *Myof* ^+/-^ heterozygous mice. **A.** Left ventricular cardiac function in male *Myof* ^+/-^ heterozygous mice (male *Myof* ^+/-^ n=9 vs. *Myof* ^+/+^ n=11). Fractional shortening (FS) (*Myof* ^+/+^ mean=69.01%, *Myof* ^+/-^ mean=46.80%); diameter, systolic (Diam, sys) (*Myof* ^+/+^ mean=0.7698mm, *Myof* ^+/-^ mean=1.408mm); diameter, diastolic (Diam, dia) (*Myof* ^+/+^ mean=2.540mm, *Myof* ^+/-^ mean=2.815mm). Mann-Whitney test was used to calculate the P value for FS due to non-normal distribution. **B.** Representative LV M-mode traces showing the difference between *Myof* ^+/-^ and *Myof* ^+/+^ control mice. **C.** Summary of electrical abnormalities seen in male *Myof* ^+/-^ mice. Representative QRS complexes on top. Representative ECG traces showing 3 cycles. **D.** Quantification of ECG parameters in male mice. Estimated QRS amplitude and duration (male *Myof* ^+/-^ n=8 vs. *Myof* ^+/+^ n=8), RR interval (male *Myof* ^+/-^ n=8 vs. *Myof* ^+/+^ n=8), J wave amplitude (male *Myof* ^+/-^ n=8 vs. *Myof* ^+/+^ n=8), QT duration (male *Myof* ^+/-^ n=4 vs. *Myof* ^+/+^ n=6, less n due to signal quality), and QTc duration (male *Myof* ^+/-^ n=4 vs. *Myof* ^+/+^ n=6, less n due to signal quality) are shown. Mann-Whitney test was used to calculate the P value for J wave amplitude due to non-normal distribution. **E.** Representative H&E staining of *Myof* ^+/-^ and *Myof* ^+/+^ cardiac tissue to assess gross morphology. **F.** Quantification of surface area in RV and LV cavities (male *Myof* ^+/-^ n=3 vs. *Myof* ^+/+^ n=3). **G.** Representative Picrosirius Red staining under polarized light microscopy of cardiac tissue to assess fibrosis burden (male *Myof* ^+/-^ n=3 vs. *Myof* ^+/+^ n=4). **H.** Quantification of Picrosirius Red signal. **I.** Pooled current-voltage (I-V) relationship from *Myof* ^+/-^ vs. *Myof* ^+/+^ isolated ventricular cardiomyocytes (cell suspension from male *Myof* ^+/-^ n=3 vs. *Myof* ^+/+^ n=3). Data are shown as mean ± SD. N was determined based on sample availability and quality of signal detected. Two-tailed unpaired Student’s t-test was used to calculate P values unless specified.

We also performed ECG recordings in 6-month-old, anesthetized mice to observe whether electrical conduction through the cardiac tissue is disrupted. *Myof* ^+/-^ male animals showed diminished QRS complex amplitude (**Fig. 8C)** along with QTc prolongation trending towards significance. The amplitude of the J wave, which is a surrogate for the T wave in humans and represents ventricular repolarization, was also reduced (**Fig. 8D**). Similar to cardiac function, electrical abnormalities were less severe in female *Myof* ^+/-^ mice, although distributed bimodally (**Fig. S8C**). We did not observe premature ventricular contractions (PVCs) at rest.

Furthermore, we assessed the structural phenotypes of the *Myof*-deficient hearts. Histological evaluation of hematoxylin & eosin (H&E)-stained tissue (**Fig. 8E**) showed significant LV dilation with a trend toward right ventricular (RV) dilation (**Fig. 8F**), recapitulating the phenotypes observed in the patient population. We also assessed fibrosis burden, a prominent arrhythmogenic phenotype in ACM. Compared to controls, *Myof* ^+/-^ mice displayed prominent fibrosis, indicating pathologic reprogramming beyond the single-cell level (**Fig. 8G-H**).

To further demonstrate that disruption of the Myof-CaV1.2 interaction has functional implications for CaV1.2 activity, we performed whole-cell patch-clamping to assess I_Ca,L_ in ventricular cardiomyocytes from *Myof* ^+/-^ compared to *Myof* ^+/+^ controls. Consistent with disrupted Ca^2+^ handling in iPSC-CM lacking MYOF, ventricular cardiomyocytes from *Myof* ^+/-^ mice showed markedly reduced I_Ca,L_ (**Fig. 8I**). Therefore, a 50% reduction of MYOF expression is sufficient to compromise the function of the CaV1.2 channel, consistent with the disrupted Myof-CaV1.2 interaction in MYOF p.G1654S carriers.

Finally, we sought to determine if *Myof* deficiency led to alterations in the expression of classic desmosomal components. Screening expression of key desmosomal proteins that harbor pathogenic variants leading to ACM (PKP2, JUP, DSP) did not show expression differences in *Myof* ^+/-^ hearts (**Fig. S9A-C**). Co-staining of MYOF with a desmosomal component, JUP, in ventricular myocytes revealed no significant co-localization (**Fig. S9D**). Together, these findings show that the cardiac-specific heterozygous *Myof* ^+/-^ knockout mouse model develops systolic dysfunction, increased fibrosis burden, and altered electrical phenotypes. Taken together with findings in cultured iPSC-CMs, we conclude that disruption of the MYOF-CaV1.2 interaction altered Ca^2+^ handling, contributed to an arrhythmogenic substrate, and recapitulated human ACM phenotypes.

## Discussion

Our work characterizes a previously undefined form of ACM driven by MYOF deficiency and a reduced interaction with CaV1.2, thus promoting arrhythmias by disrupting cardiomyocyte calcium homeostasis. These findings carry several broad implications for advancing our fundamental understanding of ACM and improving clinical diagnostic and therapeutic management strategies in this condition.

Our findings address a central unmet need in this disease, where 50–70% of ACM cases remain genetically elusive and without mechanistic explanation for pathogenesis. Anchored by our mechanistic insights into MYOF deficiency in ACM, expanding diagnostic ACM gene panels with *MYOF* variants will better enable personalized management and family screening. Moreover, although each *MYOF* variant may represent a small minority of cases, our findings indicate that MYOF deficiency collectively encompasses a much larger proportion of the general population. Specifically, we identified a family with complex ACM in which two siblings showed differential RV and LV involvement and arrhythmic burden. WES identified MYOF p.G1654S as the top candidate, segregating with disease and meeting pathogenicity criteria (**Fig. 1**). Though rare, its allele frequency suggests ∼400,000 global carriers may be at risk. Review of Penn Biobank (*10*) and BioVU data (*11*) revealed at least 3 additional patients with cardiomyopathy, ventricular arrhythmia, and ECG abnormalities, further supporting pathogenicity. Incomplete penetrance or inconsistent clinical evaluation may explain the absence of overt cardiomyopathy in some carriers, consistent with patterns seen in *PKP2* variant carriers in ACM (*29*, *30*). Gene-burden analyses in extensively phenotyped biobanks could identify additional MYOF variants and broaden our mechanistic understanding beyond p.G1654S. Nonetheless, implicating pathogenic mutations in MYOF beyond this variant and extending the relevance of MYOF deficiency to ACM, a heterozygous truncating variant p.G872QfsTer8 in MYOF was described in a patient with cardiomyopathy and arrhythmias (*8*, *9*); and another heterozygous variant p.D1575H was published in association with ARVC and SCD (*8*, *9*). Finally, emerging data have demonstrated mRNA-level *MYOF* downregulation in desmosomal models of the disease (*31*). Yet, we observed no reciprocal MYOF-dependent changes in protein expression in several key desmosomal components linked to ACM (**Fig. S9**). Such findings offer an intriguing possibility that MYOF deficiency may primarily contribute to, and potentially represent additive or synergistic pathogenic hits in, more common desmosomal ACM cases. It remains to be seen whether MYOF deficiency may also be implicated in autoantibody production against desmosomal components (*32*), another key contributor ascribed to genetically elusive ACM.

Similar to other desmosomal forms of ACM (*33*, *34*), *MYOF* variants can also exert pathogenic control through either dominant-negative (DN) or haploinsufficiency mechanisms. First, our data from patients carrying a single copy of MYOF p.G1654S or p.D1575H support a DN mechanism. Direct biochemical evidence for this mechanism comes from co-expression experiments in which MYOF^G1654S^ depleted MYOF^WT^ in a dose-dependent manner, whereas an equivalent dose of wild-type protein did not (**Fig. S4C–H**). MYOF self-associates (**Fig. S4I**), and the variant protein retains this capacity, providing a physical route by which the destabilized variant can co-recruit MYOF^WT^ to degradation and leave carriers with less MYOF than haploinsufficiency alone would predict. We also show that MYOF directly interacts with CaV1.2, and that p.G1654S disrupts this complex, impairing CaT and downstream CaV1.2 activity. This parallels an established DN mechanism in which variants in the desmosomal component *desmoglein 2* (*DSG2*) disrupt its interaction with NaV1.5 (*35*), leading to electrophysiological remodeling. Alternatively, a mechanism suggesting haploinsufficiency in a heterozygous truncating *MYOF* variant has previously been shown (*8*). Interestingly, the phenotypes observed in our haploinsufficient *Myof* ^+/-^ mouse model prove that a 50% reduction in MYOF protein expression is sufficient to drive disease. Therefore, the *Myof* ^+/-^ mouse model may represent a milder pathophenotype than a dominant-negative MYOF p.G1654S variant, where >50% of protein expression is depleted. This model of interchangeable haploinsufficient and DN mechanisms also offers a viable molecular explanation for variability in disease severity across individuals.

While ∼50% of ACM cases involve pathogenic desmosomal variants, primary Ca^2+^ mishandling has been poorly described. Our findings expand the ACM paradigm by establishing loss of MYOF-CaV1.2 interaction in p.G1654S carriers as a mechanism driving triggered activity and Ca^2+^ mishandling, with important clinical implications for antiarrhythmic therapeutic selection. Mechanistically, reduced whole-cell Ca^2+^ currents in *Myof*-deficient cardiomyocytes likely reflect diminished contractility, though may also represent a compensatory “burn-out” phase following initial Ca^2+^ overload, potentially exacerbated by structural remodeling and RYR2 de-coupling, seen in models of HFrEF (*36*, *37*). As an orthogonal mechanism, MYOF contains at least 19 predicted Ca^2+^ binding sites, suggesting it may function as a local dyadic Ca2+ buffer at the T-tubule (*38*), modulating CaV1.2 influx. NaV1.5 has also been reported to co-immunoprecipitate with MYOF (*39*), raising the possibility of additional interacting partners that could explain phenotypic heterogeneity. Overall, we identify MYOF as a critical Ca^2+^ mediator in cardiac electrophysiology, with dysregulated L-type Ca^2+^ currents representing an underappreciated and novel mechanism in ACM.

Ca^2+^ mishandling could also directly explain the systolic dysfunction in *Myof* ^+/-^ hearts, representative of the HFrEF phenotype in humans. Lower Ca^2+^ amplitude and prolonged CaT lead to decreased contractility and HFrEF development (*23*, *40*). Histologically, *Myof* ^+/-^ hearts showed prominent LV dilation, with a trend toward RV dilation, suggesting that LV dysfunction could be an initiating mechanism of pathogenesis. Chamber-specific differences may reflect environmental triggers (stress, exercise, sex) or disease stage, as biventricular dysfunction typically emerges later in ACM (*41*). MYOF may indeed play a distinct role in RV versus LV remodeling, particularly in the context of heart failure with reduced ejection fraction (HFrEF) (*42*). Specifically, MYOF is selectively upregulated in failing RVs but globally upregulated in the LV regardless of RV function. Thus, the imbalanced biventricular phenotype in *Myof* ^+/-^ mice could aid in modeling the variable RV-LV involvement seen in MYOF p.G1654S carriers. As such, further studies can now focus on studying MYOF in the context of RV vs. LV pathology.

When evaluating the electrical phenotype, we observed altered QRS complex morphology in male *Myof* ^+/-^ mice. Similar ECG findings have been documented in desmosomal mouse models, where fibrotic remodeling dampens QRS amplitude and disrupts repolarization (*43–46*). It is possible that the altered ECG signal is driven by Ca^2+^ handling dysfunction and/or is a result of fibrotic infiltrate. We have not observed an overt arrhythmia phenotype in *Myof* ^+/-^ mice, which could be explained by age effects or a lack of secondary triggers, such as stress or exercise, commonly used to induce further arrhythmogenic mouse phenotypes in ACM (*47*). Notably, iPSC-CMs from MYOF p.G1654S showed enrichment in EAD and DAD, representing the primary human phenotype.

Notably, these phenotypes showed prominent sex differences. The male *Myof* ^+/-^ model showed significantly reduced LV function and prominent fibrosis burden, similar to patient II.1 with ALVC. However, female mice did not develop severe LV dysfunction or QRS-T morphology changes (**Fig. S8**). Female *Myof* ^+/-^ data appeared bimodal, with some subgroups showing significant remodeling while others remained protected. This aligns with clinical data showing men are at higher risk of malignant arrhythmias (*48*) and SCD (*49–51*), and lower biventricular function in ACM, though this is less significant after adjusting for physical activity (*52–55*). In HFrEF, which closely resembles the male *Myof* ^+/-^ phenotype, men are more prone to chamber dilation and reduced EF, while women more commonly develop diastolic dysfunction (*56*). Together, these findings suggest that sex-based differences in ACM electrophysiology may be partly rooted in MYOF activity.

Beyond Ca^2+^ handling reprogramming, cardiac MYOF deficiency also appears to drive pathogenic processes involving cardiac structural alterations. Our RNA-seq reveals altered extracellular matrix (ECM) remodeling pathways (**Fig. 3**), a widely reported feature in ACM pathogenesis (*18*– *21*). This is consistent with the fibrosis burden we observed in the *Myof* ^+/-^ mouse model (**Fig. 8G-H**). It is possible that Ca^2+^ handling alterations could drive pro-fibrotic reprogramming in MYOF-mediated ACM, as seen in other ACM models (*57*, *58*). Calcium overload can also activate profibrotic gene programs through NFAT signaling, a transcription factor known to bind the MYOF promoter (*59*, *60*). Alternatively, a direct mechanism for MYOF in modulating the ECM could be envisioned. Fatty infiltration has been characterized in MYOF-deficient mouse skeletal muscles at 9 months of age (*5*). In cancer, MYOF promotes matrix metalloprotease (MMP) production and remodeling of the tumor microenvironment and ECM (*61*, *62*). It remains to be determined whether electrophysiological disruptions precede fibrosis and structural remodeling in MYOF-deficient hearts.

Although ACM treatment guidelines exist, diagnostic and therapeutic management requires improvement. No rigorous prospective randomized trials exist for anti-arrhythmic therapies (*63*). Βeta-adrenergic antagonists (*63*, *64*), amiodarone or sotalol, and sacubitril/valsartan (*65*) are used for the management of symptomatic arrhythmias and HF (*66*). While verapamil is not considered the first-line agent for ACM treatment, our findings implicate the efficacy of verapamil in MYOF deficiency. Verapamil may be more effective at reducing Ca^2+^ overload and EADs before LVEF declines and fibrotic remodeling occurs. This finding is consistent with a prior report describing efficacy in 50% of ARVC patients (*67*, *68*). Therefore, verapamil may be better suited for MYOF-deficient patients with high arrhythmia burden prior to structural remodeling.

Limitations to this study exist. Complementing our *in vivo* studies of *Myof* gene deletion, a *Myof ^p.G1654S/+^* mouse model could help fully elucidate the functional consequences of the dominant-negative human variant. Additionally, given the intriguing link between desmosomal ACM and MYOF deficiency, future investigation is warranted into whether MYOF interacts directly with any desmosomal components at the cellular membrane.

In conclusion, we identified a previously undefined form of non-desmosomal ACM driven by inherited MYOF deficiency. Through human molecular genetics, computational predictions, and cardiac and electrical physiology analyses in both iPSC and rodent models, we defined a mechanism of MYOF-mediated ACM Ca^2+^ mishandling via MYOF-CaV1.2 dysregulation. Consequently, this causative gene represents a candidate for developing needed targeted therapies for this deadly disease. Given the growing evidence linking MYOF to cardiomyopathy, incorporating *MYOF* to clinical cardiomyopathy gene panels would enhance genetic diagnosis and guide more precise treatment strategies for patients and clinicians.

## Materials and Methods

**Please see the Extended Methods in the Supplementary Materials.**

### Study Participants

The Institutional Review Board at the University of Pittsburgh gave ethical approval for this work (IRB MOD19050364-017). Informed consent was obtained from patients for peripheral blood monocyte collection (PBMC). Ethical approval for this study and informed consent conformed to the standards of the Declaration of Helsinki. Data from the Penn Biobank and BioVU were accessed as de-identified secondary data under each biobank’s existing institutional IRB-approved protocols. Echocardiograms, CMRs, and ECGs were over-read by multiple cardiologists blinded to pedigree and genotype. A diagnosis of ACM was made according to clinical, echocardiographic, and electrocardiographic evidence, as described by the Padua criteria (*69*).

### Exon-targeted sequencing and variant identification

Clinical grade whole genome sequencing was performed by the Stanford Medicine Clinical Genome Program with at least 40X depth using Illumina technology. Low-quality calls were excluded based on standard metrics, including read-depth, genotype quality, and allele balance.

### Inducible pluripotent stem cell reprogramming and maintenance

Peripheral blood monocytes were extracted from whole blood using SepMate™-50 density gradient centrifugation tubes per manufacturer’s protocol (StemCell, Cat# 85450) and cultured for 7 days.

### Inducible pluripotent stem cell differentiation into cardiomyocytes

iPSCs were grown to 90% confluence on Matrigel-coated 12-well plates and induced to differentiate into cardiomyocytes using the lactate selection method as described previously (*70*, *71*).

### Neonatal rat ventricular myocyte isolation

Neonatal rat ventricular myocyte (NRVM) isolation was performed as described previously (*72*).

### *Myof* ^+/-^ mouse model generation

The MYOF tm1a mice were generated by inserting loxP sites around the critical exons 6 and 7 (*73–75*). *Myof* ^tm1a/tm1a^ mice were crossed with the cardiac-specific alpha myosin-heavy chain (*Myh6*) promoter driver of Cre expression (αMyHC-Cre) (*76*) (**Fig. S8)**, generating heterozygous *Myof* ^tm1b/+;αMyHC-Cre^ (*Myof* ^+/-^) were generated and validated for loss of MYOF protein expression in the cardiac tissue. *Myof* ^tm1a/+^ (*Myof* ^+/+^) mice were used as a control.

### Statistical methods

Sample size for animal experiments was determined a priori to ensure 80% statistical power to detect a difference of at least 20% between the means of the experimental and control groups, based on an assumed standard deviation of 10%. N was also determined based on the availability of biological samples. Normality of the data was determined by Shapiro-Wilk testing and inspection of the QQ plot for linearity. Normally distributed data were analyzed by a two-tailed unpaired Student’s *t* test for comparisons between two groups; one-way ANOVA was used for comparisons between more than two groups, followed by post hoc Tukey’s analysis; two-way ANOVA was used for comparisons between more than two groups with two independent variables, followed by post hoc Tukey’s analysis. Non-normally distributed data were analyzed with Mann-Whitney nonparametric testing for comparisons between two groups; the Kruskal-Wallis test was used for comparisons between more than two groups with two independent variables, followed by post hoc Dunn’s analysis. The significance threshold was set at a *P* value less than 0.05. Data were plotted as the mean ± SD unless specified otherwise. Violin plots were reported as median ± IQR.

### List of Supplementary Materials

Extended Materials and Methods

Supplementary Tables

Table S1. Unrelated cases with cardiac involvement in MYOF p.G1654S carriers.

Table S2. Description of sgRNAs and HDR repair templates used for CRISPR/Cas9.

Table S3. Antibody information.

Table S4. Taqman primer information.

Supplementary Figures

Figure S1. Ambulatory ECG obtained from patients I.1, II.1, II.2.

Figure S2. Validation of pluripotency in iPSCs.

Figure S3. Three independent molecular Dynamics (MD) simulations of C2F WT vs. p.G1654S vs. p.D1575H structures.

Figure S4. Controls for the MYOF–CaV1.2 co-immunoprecipitation, dose-dependent depletion of MYOF^WT^ by MYOF^G1654S^, and MYOF self-association.

Figure S5. Treatment with metoprolol or dantrolene did not rescue Ca^2+^ handling defects.

Figure S6. AlphaFold modeling of the C2F domain.

Figure S7. Establishing *Myof* ^+/-^ mouse model and validating haploinsufficiency.

Figure S8. Supplementary echocardiogram and electrocardiogram information for male or female *Myof* ^+/-^ vs. *Myof* ^+/+^ mice.

Figure S9. Expression of desmosomal proteins in RV and LV tissue.

Supplemental References

## Abbreviations

ACM: Arrhythmogenic cardiomyopathy
AF: Allele frequency / AlphaFold (computational structural prediction tool)
ALT: Alternative allele
ALVC: Arrhythmogenic left ventricular cardiomyopathy
ANOVA: Analysis of variance
AP: Action potential
ARVC: Arrhythmogenic right ventricular cardiomyopathy
BioVU: Vanderbilt University Medical Center biorepository
BrdU: Bromodeoxyuridine
C2: C2 domain (Ca²⁺-binding protein domain)
CA: Concanamycin A
Ca²⁺: Calcium ion
CAD: Coronary artery disease
CaT: Calcium transient (duration)
CaV1.2: L-type voltage-gated calcium channel, alpha-1C subunit (CACNA1C-encoded)
cChange: Chromosomal change (nucleotide-level coordinate)
cDNA: Complementary deoxyribonucleic acid
CHARMM: Chemistry at Harvard Macromolecular Mechanics (force field)
Chr: Chromosome
CHX: Cycloheximide
CM: Cardiomyopathy / Cardiomyocyte (context dependent)
CMR: Cardiac magnetic resonance (imaging)
CO: Cardiac output
Co-IP: Co-immunoprecipitation
CRISPR: Clustered regularly interspaced short palindromic repeats
DAD: Delayed afterdepolarization
DAPI: 4′,6-diamidino-2-phenylindole
DEG: Differentially expressed gene
Diam, dia: Diameter, diastolic
Diam, sys: Diameter, systolic
DMEM: Dulbecco’s Modified Eagle Medium
DMSO: Dimethyl sulfoxide
DN: Dominant negative
DNA: Deoxyribonucleic acid
dNTP: Deoxynucleotide triphosphate
DPBS: Dulbecco’s phosphate-buffered saline
DSG2: Desmoglein 2
DSP: Desmoplakin
dsRNAbd: Double-stranded RNA binding domain
dTTP: Deoxythymidine triphosphate
dUTP: Deoxyuridine triphosphate
EAD: Early afterdepolarization
ECG: Electrocardiogram
ECM: Extracellular matrix
EDTA: Ethylenediaminetetraacetic acid
EF: Ejection fraction
EHD2: EH-domain containing 2
EPC: Erythroid progenitor cell
FAT1: FAT atypical cadherin 1
FDA: United States Food and Drug Administration
FPKM: Fragments per kilobase of transcript per million mapped reads
FS: Fractional shortening
GAPDH: Glyceraldehyde 3-phosphate dehydrogenase
gnomAD: Genome Aggregation Database
GO: Gene ontology
HA: Hemagglutinin (epitope tag)
HDR: Homology-directed repair
H&E: Hematoxylin and eosin
HE: Heterozygous
HEK293T: Human embryonic kidney 293T cell line
HF: Heart failure
HFrEF: Heart failure with reduced ejection fraction
HO: Homozygous
HSP90: Heat shock protein 90
IB: Immunoblot
ICa,L: L-type calcium current
ICD: Implantable cardioverter-defibrillator
ID: Identification number
IF: Immunofluorescence
IP: Immunoprecipitation
iPSC: Induced pluripotent stem cell
iPSC-CM: Induced pluripotent stem cell-derived cardiomyocyte
IQ domain: Isoleucine-glutamine domain (calmodulin-binding motif)
IQR: Interquartile range
JUP: Junction plakoglobin
KO: Knockout
LGE: Late gadolinium enhancement
LV: Left ventricle
LVAD: Left ventricular assist device
LVAW, dia: Left ventricular anterior wall, diastolic
LVAW, sys: Left ventricular anterior wall, systolic
LVEDV: Left ventricular end-diastolic volume
LVEF: Left ventricular ejection fraction
LVESV: Left ventricular end-systolic volume
LV mass, corr: Left ventricular mass, corrected
LVPW, dia: Left ventricular posterior wall, diastolic
LVPW, sys: Left ventricular posterior wall, systolic
MAF: Mean (minor) allele frequency
MD / MDS: Molecular dynamics / molecular dynamics simulations
MF: Molecular function
MMP: Matrix metalloprotease
Myh6 / αMyHC: Alpha myosin heavy chain (gene/protein, mouse cardiac-specific promoter)
MYOF: Myoferlin
NaV1.5: Voltage-gated sodium channel 1.5 (SCN5A-encoded)
NFAT: Nuclear factor of activated T cells
NRVM: Neonatal rat ventricular myocyte
PAM: Protospacer-adjacent motif
PBMC: Peripheral blood mononuclear cell
PBS: Phosphate-buffered saline
pChange: Protein change (amino acid substitution coordinate)
PCR: Polymerase chain reaction
Penn / PMBB: University of Pennsylvania / Penn Medicine BioBank
PKP2: Plakophilin 2
PLA: Proximity ligation assay
P/LP: Pathogenic / likely pathogenic (variant classification)
PVC: Premature ventricular contraction (complex)
REF: Reference allele
REVEL: Rare exome variant ensemble learner
RMSD: Root-mean-square deviation
RNA: Ribonucleic acid
RNA-seq: RNA sequencing
RNP: Ribonucleoprotein complex
RT-qPCR: Quantitative reverse-transcription polymerase chain reaction
RV: Right ventricle / right ventricular
RVEF: Right ventricular ejection fraction
RYR2: Ryanodine receptor 2
SCD: Sudden cardiac death
SD: Standard deviation
SDS-PAGE: Sodium dodecyl sulfate–polyacrylamide gel electrophoresis
sgRNA: Single-guide RNA
siMYOF: Myoferlin-targeting small interfering RNA
siNC: Scrambled (negative control) small interfering RNA
siRNA: Small interfering RNA
SR: Sarcoplasmic reticulum
SV: Stroke volume
TWI: T-wave inversion
UPMC: University of Pittsburgh Medical Center
Vol, dia: Volume, diastolic
Vol, sys: Volume, systolic
VT: Ventricular tachycardia
VUS: Variant of uncertain significance
WES: Whole-exome sequencing
WT: Wild type

## Data Availability

All data are available in the main text or the supplementary materials. RNA sequencing data have been deposited in a public repository and will be made available upon publication. GEO accession: GSE347506. Plasmids and the Myof mouse model generated in this study are available from the corresponding author under a material transfer agreement (MTA).

## Acknowledgments

This work was supported by the WoodNext Foundation and the American Heart Association SFRN (to S.Y.C.), National Institutes of Health grant 1F30HL170649-01 (A. K.). Work performed in the UPMC Hillman Cancer Center Tissue and Research Pathology Services (TARPS) Shared Resource Facility, and services and instruments used in this project were graciously supported, in part, by the University of Pittsburgh and the National Cancer Institute of the National Institutes of Health under Award Number P30CA047904. Work performed in the Pitt Biospecimen Core (RRID: SCR_025229) and services and instruments used in this project were supported, in part, by the University of Pittsburgh, the Office of the Senior Vice Chancellor for Health Sciences.

## Author contributions

Conceptualization: AK, MA, SYC

Methodology: AK, MA, IC, GS, HW

Software: AIM, YS, SO, CJC

Formal analysis: AK, MA, AIM, YS, SO, CJC, SMN

Investigation: AK, MA, IC, AlK, NK, WSG, YYA, YY, SJ, RJR, YT, CF, MDC

Resources: MA, DYZ, CR, JSA, JP, ELB, SAS, JRM, GS, HW, MZ, PJK, MSG, VNP, SYC

Data curation: AK, MA, AIM, SO

Writing – original draft: AK, MA, SYC

Writing – review & editing: AK, MA, VNP, MSG, GS, MZ, SMN, SYC

Visualization: AK, YS, AIM

Supervision: SYC, VNP, MSG, GS, MZ

Funding acquisition: AK, SYC

Project administration: AK, SYC

