## Supplemental Information for "Pathogenic variants in myoferlin (*MYOF)* cause arrhythmogenic cardiomyopathy"

### Extended Materials and Methods

#### Study Participants

The Institutional Review Board at the University of Pittsburgh gave ethical approval for this work (IRB MOD19050364-017). Informed consent was obtained from patients for peripheral blood monocyte collection (PBMC). Ethical approval for this study and informed consent conformed to the standards of the Declaration of Helsinki. Echocardiograms (cardiac magnetic resonance imaging), CMRs, and electrocardiograms (ECGs) were over-read by multiple cardiologists blinded to pedigree and genotype. A diagnosis of ACM was made according to clinical, echocardiographic, and electrocardiographic criteria as described by the Padua criteria (*1*).

#### Exon-targeted sequencing and variant identification

Clinical grade whole genome sequencing was performed by the Stanford Medicine Clinical Genome Program with at least 40X depth using Illumina technology. Low-quality calls were excluded based on standard metrics, including read depth, genotype quality, and allele balance. High-confidence variants were subsequently annotated using population frequency data from gnomAD (32461654) (*2*), functional impact scores from REVEL (27666373) (*3*), and clinical significance from ClinVar (29165669) (*4*). For segregation analysis, each variant's two alleles were recorded as binary indicators, "1" for the presence of the minor allele and "0" for the reference allele. Variants were prioritized based on the following criteria: 1. The variant is present on at least one allele (e.g., "1_0", "0_1", or "1_1") in both siblings but absent on both alleles in the parent I.2 (i.e., "0_0"), 2. The variant results in a missense amino acid substitution. 3) The minor allele frequency (MAF) is below 0.5% in all populations reported in gnomAD. And 4) The REVEL score exceeds 0.6, indicating a high likelihood of pathogenicity.

### DNA extraction and Sanger sequencing

PBMCs were collected from primary patient samples. DNA was extracted using QuickExtract (Biosearch Technologies) per the manufacturer's protocol. Briefly, DNA was mixed in QuickExtract and heated at 65 °C for 6min. 5 μL was subsequently used for PCR amplification using Q5 High-Fidelity Master Mix (NEB). The following amplification primers were used 5'-TGACACCTTTACCCGGGATG-3' and 5'AAGCAGCTGTGTTGGTCTCA-3'. Amplified PCR product was sequenced using Sanger sequencing technology at a core facility (Azenta).

### Clinical summary of a family with ARVC and ALVC

We identified a family with variable presentation of arrhythmogenic cardiomyopathy (**Fig. 1A**). Clinical genetic testing failed to reveal any pathogenic variants in established cardiomyopathy-associated genes. Two affected siblings have differential ventricle involvement and arrhythmia burden: with one fulfilling criteria for ARVC (II.2), and the other for ALVC (II.1). Their parent (I.1, deceased) had non-ischemic cardiomyopathy with ventricular arrhythmias. Parent I.1 had a significant history of familial sudden cardiac death (SCD). Two relatives died suddenly. Parent I. 2's side is unaffected from a cardiomyopathy standpoint.

### II.1

Proband II.1 first became known to the general cardiology clinic for evaluation for cardiomyopathy due to a longstanding history of premature ventricular contractions (PVCs). At the first presentation, the CMR findings were solely remarkable for mildly depressed left ventricular ejection fraction (LVEF 52%) and ring-like diffuse subepicardial and mid-wall late gadolinium enhancement (LGE), in the LV myocardium. Following a presyncopal episode, the proband underwent a repeat CMR, which showed worsened LV function (LVEF 46%) with global hypokinesis and moderate LV dilation. Subepicardial LGE was redemonstrated on this study (**Fig. 1B**). Ambulatory monitoring revealed a high burden of PVCs. RV biopsy showed myocyte hypertrophy without evidence of acute inflammation, sarcoidosis, iron overload, or other storage disorder. Electrocardiogram (ECG) demonstrated sinus bradycardia, borderline nonspecific interventricular conduction delay, and no T-wave inversions (TWI) (**Fig. S1**). In the setting of idiopathic LV dilation and dysfunction, ring-like LGE is pathognomonic for arrhythmogenic cardiomyopathy of the LV, high PVC burden, and unexplained syncope. The patient underwent an electrophysiology study, including extrastimulus, resulting in ventricular fibrillation. An implantable cardiac defibrillator was placed for primary prevention of SCD. Clinical genetic testing of the proband identified a variant of uncertain significance (VUS) p.Pro119Leu in Plakoglobin (JUP) that is common in the general population (rs376123010, gnomAD 0.002%) (*2*). This VUS has not been reported in the literature in individuals affected by ARVC.

**II.2**

Patient II.2 was evaluated for cardiomyopathy in light of sibling's (II.1) diagnosis and history of presyncope and ventricular bigeminy. The cardiac CMR showed a dilated RV (RVEF = 58%) with aneurysms and dyskinesis in the mid RV free wall. Atypical LGE in the LV inferolateral wall was noted (**Fig. 1B**). The LV was mildly dilated but with normal systolic function (LVEF = 61%). A PVC burden of 18.7% was identified on ambulatory monitoring, along with 8 ventricular triplets. Signal-averaged ECG revealed a filtered QRS duration of 118ms, a root mean square voltage of the last 40 ms of 25 mV, and a signal duration less than 40 μV of 31ms (**Fig. S1)**. ARVC diagnosis was established based on (1) major imaging criteria (RV size and aneurysms), and 2 minor criteria (>500 PVCs in a 24-hour period and filtered QRS duration > 114ms). After shared decision-making based on available risk calculators (*5*), a primary prevention ICD was implanted.

**I.1**

Patient I.1 suffered from VF arrest. The patient had a history of PVCs, atrial fibrillation, and nonischemic cardiomyopathy. The CMR studies were negative for evidence of ARVC but showed complete LV transmural LGE of the basal to mid inferolateral, basal inferior, and portion of the basal anterolateral wall segments, as well as depressed LVEF<40% (not shown). The records indicated a scar pattern consistent with myocardial infarction; however, no acute myocardial infarction was registered (**Fig. S1**). Cardiac catheterization did not reveal any obstructive coronary artery disease.

**I.2**

Parent I.2 has a history of atrial fibrillation, sick sinus syndrome, and mitral valve disease. Patient does not have a family history of cardiomyopathy.

**Clinical summary of a case found in Penn Biobank**

We were able to identify a patient with a diagnosis of non-ischemic cardiomyopathy and ventricular tachycardia in the Penn Biobank. The patient underwent a heart transplant. Family history was notable for SCD. The last echocardiogram prior to transplant showed a severely dilated LV (LV end-diastolic diameter = 10.0 cm); normal wall thickness; global hypokinesis with minimal segmental variation; akinetic LV apex. LV EF was severely decreased, estimated at 10%.

The RV was mildly dilated with moderately reduced systolic function; Tricuspid Annular Plane Systolic Excursion (TAPSE) = 1.5 cm (reduced). Increased LV trabecular pattern was seen along the posterior wall, but not in the classic pattern for noncompaction cardiomyopathy. Before transplant, the patient received a left ventricular assist device (LVAD) and an ICD. The last documented ICD interrogation revealed numerous NSVT and VT episodes. Post-transplant, cardiac pathology showed biventricular cardiomyocyte hypertrophy and patchy fibrous thickening of the endocardium with no evidence of iron, amyloid, or glycogen deposition. ECG revealed normal sinus rhythm with sinus arrhythmia; right axis deviation; ST & T wave abnormality (consistent with inferolateral ischemia); nonspecific intraventricular conduction delay; prolonged QT.

**Evaluation of P/LP variants in Penn Biobank and BioVU**

Data from the Penn Biobank and BioVU were accessed as de-identified secondary data under each biobank's existing institutional IRB-approved protocols. A total of 169 genes from the PreventionGenetics cardiomyopathy panel (Exact Sciences) were analyzed. ClinVar (*2*) was queried for all variants with a 2-star rating using the most recent ClinVar annotations available from their FTP site (release date: 2025-05-04). Variants with conflicting interpretations were excluded. Remaining variants were filtered to include only those classified as pathogenic or likely pathogenic (P/LP), yielding 7,776 variants. For each P/LP variant, the Penn Medicine BioBank (PMBB) was searched using both exome and imputed genotype data. This process identified 1,080 variants present in the dataset. For each identified variant, ClinVar ID, gene, pathogenicity, and genotype status for the *MYOF* variant carrier were documented. The patient under study did not carry any of these 1,080 variants.

Patient data were derived from Vanderbilt University Medical Center's Synthetic Derivative (SD)(*6*), a deidentified electronic health record (EHR) database containing longitudinal clinical data between 1995 and 2025. The EHR conformed to the Observational Medical Outcomes Partnership (OMOP) Common Data Model (CDM). A complete history of patient notes, diagnosis codes, procedures, medications, measurements, and visits was extracted for each patient.

**Inducible pluripotent stem cell (iPSC) reprogramming and maintenance**

Peripheral blood monocytes were extracted from whole blood using SepMate™-50 density gradient centrifugation tubes per manufacturer's protocol (StemCell, Cat# 85450) and cultured for 7 days. Enriched erythroid progenitor cells were used for reprogramming using the CytoTune™-iPS 2.0 Sendai Reprogramming Kit (ThermoFischer, Cat#A16517) per manufacturer's protocol. Briefly, the Yamanaka reprogramming factors, Oct, Sox2, Klf4, and c-Myc were transiently transduced into $5x10^5$ EPCs and maintained in StemPro™-34 medium without cytokines (ThermoFischer, Cat#10639011) for 7 days and transitioned to mTeSR1 media (StemCell Cat# 85850). Emerging iPSC clones were live-stained with Alexa 488-Tra-1-60 conjugated antibodies (ThermoFisher, Cat# A25618) and manually isolated. Stable iPSC cultures were passaged at least 10-20 times before pluripotency characterization and differentiation.

Reprogrammed iPSCs were maintained in mTeSR1 media (StemCell Cat# 85850) on Matrigel-coated plates (Corning Cat#354277). Cells were passaged 4-5 days after reaching 80% confluency using a gentle cell dissociation reagent (StemCell Cat# 100-0485) and plated in medium supplemented with Y-27632 Dihydrochloride (StemCell Cat#72304). After 24hrs, the medium was changed without Y-27632 Dihydrochloride and subsequently changed daily. Pluripotency of iPSC lines was confirmed using immunofluorescent staining and gene expression**.** To further assess for pluripotency, iPSCs were differentiated into 3 germ layers (StemCell Cat#05230). iPSC from all patient lines differentiated into ectoderm (Nestin), mesoderm (Brachyury), and endoderm (SOX17), which was confirmed using immunofluorescent staining. Refer to **Table S3-4** for antibody and Taqman primer information.

**Inducible pluripotent stem cell differentiation into cardiomyocytes**

iPSCs were grown to 90% confluence on Matrigel-coated 12-well plates and induced to differentiate into cardiomyocytes using the lactate selection method as described previously(*7*, *8*). Briefly, iPSCs were rinsed with DPBS (ThermoFischer) and incubated with 5 μM CHIR99021 (StemCell Cat#72052) diluted in PRMI 1640 (Gibco, Cat# 11875093) supplemented with B27 minus insulin (Gibco, Cat#A1895601). After 48hrs, the media was changed to 5uM IWR-endo (StemCell, Cat#72562) and incubated for 48hrs. Subsequently, the media was changed daily with RPMI 1640+B27 minus insulin for 2 days, followed by 3 days of culture with RPMI 1640 + B27 complete (Gibco, Cat# 17504044). Cells were maintained in RPMI 1640, no glucose (Gibco, Cat# 11879020) with B27 complete and sodium lactate until day 13 of differentiation. On day 13, cells were dissociated using Accutase (StemCell, Cat#07920) and plated back. Cells were maintained in lactate-supplemented medium until day 30-40. All experiments were performed using day 30-35 cells.

**CRISPR/Cas9 editing**

Small-guide RNA (sgRNA) and homology repair (HDR) templates targeting rescue c.4960A>G, knock-in c.4960G>A, and knock-in c.4723G >C were designed using the online CRISPR design tool Benchling (https://benchling.com). Ribonuclear complex (RNPs) were assembled by incubating sgRNA (Integrated DNA Technologies) and recombinant Cas9 protein (New England Biosciences) in Opti-MEM (Gibco, Cat# 31985070), followed by the addition of HDR templates. iPSCs were dissociated and resuspended in R buffer (Invitrogen, Catalog#MPK10096). RNPs and HDR were electroporated (1650V, 10msec, 2 pulses) into $10^6$ dissociated iPSCs via Neon Electroporation System (Invitrogen) and Neon Transfection system 100uL kit (Invitrogen, Catalog#MPK10096). Cells were seeded in 6-well Matrigel-coated plates in mTeSR1 supplemented with Y-27632. Y-27632 was removed from the medium after 24hrs. Surviving clones were manually isolated and genotyped. Refer to **Table S2** for sequence and Cas9 information.

**High-throughput RNA sequencing and analysis**

iPSC-CM were lysed in Qiazol (Qiagen). RNA extraction was performed using the Rneasy Mini Kit (Qiagen) per the manufacturer's protocol.

*Sample quality control*

RNA integrity was assessed using the Bioanalyzer 2100 system (Agilent Technologies, CA, USA).

*Library preparation for Transcriptome sequencing*

Non strand specific library Messenger RNA was purified from total RNA using poly-T oligo-attached magnetic beads. After fragmentation, the first strand cDNA was synthesized using random hexamer primers, followed by the second strand cDNA synthesis. The library was ready after end repair, A-tailing, adapter ligation, size selection, amplification, and purification. The library was checked with Qubit and real-time PCR for quantification and a bioanalyzer for size distribution detection.

*Strand-specific library*

Messenger RNA was purified from total RNA using poly-T oligo-attached magnetic beads. After fragmentation, the first strand cDNA was synthesized using random hexamer primers. Then the second strand cDNA was synthesized using dUTP, instead of dTTP. The directional library was ready after end repair, A-tailing, adapter ligation, size selection, amplification, and purification. The library was checked with Qubit and real-time PCR for quantification and a bioanalyzer for size distribution detection.

*Clustering and sequencing*

After library quality control, different libraries were pooled based on the effective concentration and targeted data amount, then subjected to Illumina sequencing. The basic principle of sequencing is "Sequencing by Synthesis", where fluorescently labeled dNTPs, DNA polymerase, and adapter primers are added to the sequencing flow cell for amplification. As each sequencing cluster extends its complementary strand, the addition of each fluorescently labeled dNTP releases a corresponding fluorescence signal. The sequencer captures these fluorescence signals and converts them into sequencing peaks through computer software, thereby obtaining the sequence information of the target fragment.

*Data quality control*

Raw data (raw reads) of fastq format were first processed through in-house Perl scripts. In this step, clean data (clean reads) were obtained by removing reads containing adapters, reads containing poly-N, and low-quality reads from raw data. At the same time, Q20, Q30, and GC content of the clean data were calculated. All the downstream analyses were based on the clean data with high quality.

*Quantification of gene expression level*

FeatureCounts v1.5.0-p3 was used to count the number of reads mapped to each gene. And then FPKM of each gene was calculated based on the length of the gene and the read count mapped to this gene. FPKM, expected number of Fragments Per Kilobase of transcript sequence per Millions base pairs sequenced, considers the effect of sequencing depth and gene length for the read count at the same time, and is currently the most commonly used method for estimating gene expression levels.

*Differential expression analysis*

Differential expression analysis was performed using the DESeq2 R package(1.20.0). DESeq2 provides statistical programs for determining differential expression in digital gene expression data using models based on the negative binomial distribution. The resulting P-value is adjusted using Benjamini and Hochberg's methods to control the error discovery rate. The corrected P-value $\leq$ 0.05 & $|\log2\ (\text{foldchange})| \geq 1$ was set as the threshold of significant differential expression.

**Protein Structure and Analysis**

The predicted Alphafold structure of human myoferlin used in this paper was downloaded from UniProt Q9NZM1. Mutants were generated using Pymol. Docking models and computational free energies were obtained using ClusPro (*9*) and FastContact (*10*), respectively.

**Simulation Protocol**

Molecular dynamics simulations (MDS) were run for 300 ns with NAMD 2.14 for Linux-x86_64-multicore-CUDA (*11*) using CHARMM 36 jul24 force field (*12*, *13*). The first 150 ns of each run were discarded as equilibration time. We used VMD (*14*) 1.9.4a53 to center each protein in a cuboid TIP3P water box with a 10 Å distance from the protein surface to the box edges and closeness parameter of 2.4 Å. The system was neutralized, and 0.15 mol/L NaCl was added using the VMD autoionize tool. Pressure (Langevin piston) was set at 1 bar with piston period = 100, and piston decay = 50. The non-bonded (direct) interaction cutoff distance was set to 12 Å, and time steps of 2 fs.

The simulated structures were prepared as follows. First, we performed Langevin dynamics with a damping (friction) coefficient of 1/ps, and harmonic constraints with a scaling coefficient of 0.8 in all backbone atoms excluding hydrogens. The starting temperature was 60 K and increased by 60 K after 500 timesteps up to 310 K. Velocities were re-initialized from a Maxwell-Boltzmann distribution each time the temperature was reset. The protein structure was then run for another 50 ps, followed by a 0.5 ns run without harmonic constraints.

**Quantitative RT-qPCR**

Cells were lysed in Qiazol (Qiagen), and RNA extraction was performed using Rneasy Mini Kit (Qiagen) per the manufacturer's protocol. 500ng of RNA was reverse transcribed to complementary DNA (cDNA) using the High Capacity cDNA reverse transcription kit (ThermoFiser, Cat# 4368813). cDNA samples were diluted 1:30 with $H_2O$ and mixed with TaqMan primers. Quantitative RT-PCR (RT-qPCR) was performed using the Applied Biosystems QuantStudio 6 Flex Fast Real Time PCR system. Relative fold change ($2^{-\Delta\Delta Ct}$) of each transcript was normalized to *GAPDH*. **Table S4** for antibody and Taqman primer information.

**Immunoblotting**

iPSC-CM monolayer was rinsed with ice-cold PBS and harvested in 1.5% Triton-X in PBS supplemented with protease inhibitor (ThermoFisher). Quantification of the soluble protein concentration was performed by the BCA assay (ThermoFisher, Cat#23250). For reducing and denaturing gels, protein (20ug) was loaded into a 4-15% gradient SDS-PAGE gel (Biorad) and allowed to separate for 1.5hrs at 110V in 1x Tris/glycine/SDS buffer. Protein was transferred onto nitrocellulose membrane (Biorad, Cat#1620115) overnight at 40mA 1x Tris/glycine buffer. Membrane was blocked in 5% blotting grade milk (Biorad) diluted in phosphate buffered saline (PBS) for 1hr at room temperature while rocking and incubated with primary antibodies overnight at 4 °C. After washes with 0.01% TritonX-100 diluted in PBS, membranes were incubated with anti-rabbit or anti-mouse (1:2000) secondary horse protein radish (HRP) conjugated antibodies (Dako) diluted in 5% blotting grade milk for 1hr at room temperature. Secondary antibodies conjugated to HRP (anti-mouse or anti-rabbit, as appropriate) were applied, and signals were detected using enhanced chemiluminescence (ECL) reagents (Thermo Fisher Scientific) and imaged using Biorad ChemiDoc XRS+ and ImageLab 6.0.1 software. Band intensity quantification was performed using ImageLab 6.0.1 software.

**Overexpression of MYOF in HEK293T cells**

HEK293T cells were cultured in Dulbecco's Modified Eagle Medium (ThermoFisher, Cat#12634010) supplemented with 10% fetal bovine serum (FBS, Sigma). Cells were seeded into 100 mm tissue culture dishes to achieve ~60% confluency at the time of transfection. Cells were transfected with Plasmids encoding $MYOF^{WT–FLAG}$ or $MYOF^{G1654S–FLAG}$ (Origene, Cat#RC218766) using Lipofectamine 2000 per manufacturer's protocol (ThermoFisher). After 24hrs of transfection, protein lysates were harvested for non-reducing gel analyses. For degradation inhibition experiments, HEK293T cells were transfected with expression plasmids for 24hrs and subsequently treated with 100nM epoxomicin (Adooq, A12730) or 100nM Concanamycin A (MCE, HY-N1724) for another 24hrs.

**Site Directed Mutagenesis**

To establish the $MYOF^{G1654S–FLAG}$ expression plasmid, site directed mutagenesis was performed using QuickChange II XL Site Directed Mutagenesis Kit (Agilent, Cat#200521). Briefly, $MYOF^{WT–FLAG}$ (Origene, Cat#RC218766) was PCR amplified using 5'-ctcctctggtatgctgcagtgggacccaa-3' and 5'- ttgggtcccactgcagcataccagaggag-3'. The PCR product was transformed into XL 10-Gold Ultracompetent cells per the manufacturer's protocol.

**HA-tagged MYOF constructs, co-expression assay, and MYOF self-association**

$MYOF^{WT–HA}$ was obtained commercially (**Addgene, Cat#22443**), and $MYOFG^{1654S–HA}$ was generated from this construct by site-directed mutagenesis using the QuikChange II XL Site-

Directed Mutagenesis Kit (Agilent, Cat#200521) with the primers described above, and verified by Sanger sequencing. For co-expression assays, HEK293T cells were seeded in 6-well plates to ~60% confluency and co-transfected with Lipofectamine 2000 (ThermoFisher) using a fixed amount of one MYOF construct together with a graded amount (0, 125, 500, 1000, and 2000 ng) of the second, differentially tagged construct, as indicated in **Fig. S4**. Lysates were harvested 48 hours after transfection in 1.5% Triton X-100 lysis buffer supplemented with protease inhibitors, resolved on 6% SDS-PAGE gels, and immunoblotted for HA, FLAG, and HSP90. Band intensities were quantified in ImageJ and normalized to HSP90. To assess MYOF self-association, HEK293T cells were co-transfected with the indicated combinations of FLAG- and HA-tagged $MYOF^{WT}$ and $MYOF^{G1654S}$ constructs, and immunoprecipitation with anti-FLAG M2 magnetic beads (Sigma-Aldrich) was performed as described above for the MYOF–CaV1.2 co-immunoprecipitation. Immunoprecipitates, inputs, and post-immunoprecipitation supernatants were resolved on 6% SDS-PAGE gels and immunoblotted with an anti-HA antibody. Refer to **Table S3** for antibody information.

**Cycloheximide CHX chase assays**

HEK293T cells were cultured and transfected with 0.5 µg of $MYOF^{WT}$ or $MYOF^{G1654S}$ expression plasmids overnight. The following day, lysates were harvested in 1.5% TritonX-100 lysis buffer supplemented with protease inhibitor. Cells were treated with 300ug/mL cycloheximide (CHX) (Sigma) at t=0. Lysates were subsequently harvested at t=2, t=4, and t=6. HSP90 was selected as a loading control based on its superior stability post-CHX treatment (compared to beta-ACTIN and GAPDH). Half-life was quantified by fitting an exponential decay equation model to the $MYOF^{WT}$ and $MYOF^{G1654S}$ curves.

**Optical measurements of live calcium transients**

Measurement of live calcium transients was performed as described previously (*15*)**.** At day 20 of iPSC-cardiomyocyte differentiation, cells were dissociated with Accutase (Gibco) and plated at $1.25x10^5$cells/plate on 35mm glass-bottom plates (Mattek), coated with Matrigel (Corning). Cells were cultured until day 30-35 before live calcium acquisition. For NRVM image acquisition, cells were plated at $2x10^5$ density on gelatin (Sigma, Cat# G9136) coated 35mm cell culture imaging dishes (ibidi, Cat# 81156). Image acquisition was performed using the Nikon Ti-2E microscope platform enhanced with SPECTRA III Light Engine solid-state LED light source and Hamamatus Orca-fusion Gen-III sCMOS camera. Videos were acquired at 2048 × 2048 resolution and a minimum of 25 frames per second. For ratiometric recordings, cells were loaded with 5 μM Fura-2 AM for 20min at 37 °C and allowed to recover before image acquisition at 37 °C. Imaging dishes were mounted and paced at 1Hz (10 volts/cm, bipolar pulse, 10ms wave width) using a field stimulation (Warner Instrument LLC, Cat#RC-21BRFS). Calcium signals were pre-processed using Nikon NIS software and analyzed further using a customized Python algorithm.

**Optical measurement of live action potentials**

Measurement of live action potentials was performed as described previously (*16*). Cells were loaded with 0.1% volume Fluovolt membrane potential dye (Invitrogen, Cat# F10488) and incubated at 37 °C for 20min. Cells were washed twice and allowed to recover before live image acquisition. Image acquisition was performed using the Nikon Ti-2E microscope platform enhanced with SPECTRA III Light Engine solid-state LED light source (490nm excitation). Videos were acquired at 2048 × 2048 resolution and a minimum 25 frames per second.

**Neonatal rat ventricular myocyte isolation**

Neonatal rat ventricular myocyte (NRVM) isolation was performed as described previously (*17*). Briefly, 1–2-day-old Sprague-Dawley rat pups were anaesthetized with 5% isoflurane until unresponsive to pain stimuli. Ventricular tissue was quickly excised and washed 3 times in ice cold KH buffer titrated to pH 7.5 (NaCl 140 mM, KC1 4.8 mM, $MgSO_4$-$7H_2O$ 1.2 mM, dextrose 12.5 mM, $NaHCO_3$ 4 mM, $NaH_2PO_4$ 1.2 mM, and HEPES 10 mM). Tissue was cut into 1-2mm pieces and dissociated in trypsin (0.04%) (Gibco, Cat#15-090-046) and collagenase (0.4mg/ml) (Worthington, Cat# LS004176) at 37 °C while stirring. Digestion was terminated with 10% fetal bovine serum (FBS) in DMEM (Cat# 11965092). Cell suspension was pre-incubated for 90min to allow attachment of cardiac fibroblasts. Media was supplemented with 0.1M bromodeoxyuridine (BrdU) to halt fibroblast proliferation. After 24hrs of culture, the growth media was changed to DMEM supplemented with 0.1% Insulin-transferrin-selenium (Gibco, Cat# 51500056). NRVMs were used for experiments 24hrs after culturing.

**Silencer RNA knockdown in NRVMs**

Transient transfections were performed in TransIT-TKO Transfection Reagent.

Transient knockdown of MYOF in NRVMs was performed with 50nM pooled Silencer.

Select siRNAs (Thermo Fischer, Cat# 4390771; ID: s159816, s159817, s159815) or 50nM negative control silencer (Thermo Fischer, Cat# 4390843). Additional media was re-supplemented after 24hrs. Experiments were performed 72hrs post transfection.

**Isolation of cardiac tissue for immunofluorescence**

Cardiac tissue was isolated as previously described (*18*). Briefly, cardiac tissue was perfused with PBS followed by 2% paraformaldehyde (PFA). Tissue was fixed in 2% PFA for 2hrs at 4 °C, followed by 24hr incubation in 30% sucrose at 4 °C. Tissue was then snap frozen in methyl butane followed by liquid nitrogen.

**Immunostaining and confocal microscopy**

Cells were cultured in Matrigel or collagen-coated Lab-Tek II chamber slides (Thermo). Slides were washed phosphate buffered saline (PBS) and cells fixed with 2% paraformaldehyde for 15 min. Cells were permeabilized for 1 hr in 0.1% Triton X-100 diluted in PBS and blocked for 1 hr with 5% normal goat serum and 2% bovine serum albumin diluted in PBS. Primary antibodies were incubated overnight at 4°C, diluted in blocking buffer**.** The next day, slides were washed and incubated in 1% BSA in PBS with secondary antibodies (1:2000), incubated at room temperature (RT) for 1.5 hrs protected from light. Samples were washed and mounted with DAPI (Thermo Fischer, Cat# P36935). Images were acquired on an Olympus Fluoview 300 confocal microscope with a 60x oil immersion objective.

For tissue staining, cryo-sectioned (7um) mouse cardiac samples were washed twice with ice-cold PBS. Tissues were permeabilized with 0.1% Triton X-100 in PBS for 10min and blocked in 5% normal goat serum and 2% bovine serum albumin diluted in PBS for 1hr at RT. For mouse-on-mouse staining, mouse IgG blocking serum (Thermo, Cat# R37621) was used for 1hr at RT. After washing with 1% BSA, cells were incubated overnight at 4°C with primary antibodies diluted in blocking buffer. The next day, slides were washed and incubated with secondary antibodies (1:2000) diluted in blocking buffer for 1hr at RT. Samples were washed and mounted with DAPI.

Images were acquired on an Olympus Fluoview 300 confocal microscope with a 60x oil immersion objective. Refer to **Table S3** for antibody information.

**Co-immunoprecipitation**

To investigate whether MYOF directly interacts with the L-type calcium channel CaV1.2, co-immunoprecipitation (Co-IP) assays were performed using transiently transfected HEK293T cells. Both wild-type ($MYOF^{WT}$) and mutant ($MYOF^{G1654S}$) constructs tagged with a C-terminal FLAG epitope were used for expression, along with an HA-tagged full-length α-subunit of CaV1.2. After 48 hours, cells were washed with PBS and lysed in 1000 µL of Triton X-based lysis buffer (50 mM Tris-HCl pH 7.5, 160 mM NaCl, 1.5% Triton X-100, 1 mM EDTA) supplemented with protease and phosphatase inhibitors (Roche). Lysates were incubated on ice for 30 minutes and centrifuged at 14,000 rpm for 15 minutes at 4°C to remove debris. Supernatants were transferred to fresh tubes, and 100 µL of each was saved as pre-IP input and stored at −80°C. The remaining lysates were incubated with anti-FLAG M2 magnetic beads (Sigma-Aldrich) overnight at 4°C with gentle rotation. Beads were washed twice with cold TBS and once with lysis buffer to remove non-specific interactions before elution with 4× Laemmli sample buffer at 95°C for 5 minutes. Samples were resolved on 6% SDS-PAGE gels and analyzed by western blotting as described. Inputs (10%) were loaded alongside IP samples to confirm protein expression levels. Refer to **Table S3** for antibody information.

**Dose-dependent depletion**

HEK293T cells were co-transfected with the indicated combinations of FLAG- and HA-tagged $MYOF^{WT}$ and $MYOF^{G1654S}$ constructs. HEK293T cells were co-transfected with differentially epitope-tagged MYOF expression constructs and harvested 48 hours later. Complexes were captured with anti-FLAG magnetic beads and immunoblotted for HA. For each condition, the immunoprecipitate (IP), the whole-cell lysate before immunoprecipitation (input), and the unbound fraction remaining after immunoprecipitation (post-IP) were loaded. The wild-type/variant pair was tested in both tag orientations ($MYOF^{WT–FLAG}$ with $MYOF^{G1654S–HA}$, and $MYOFG^{1654S–FLAG}$ with $MYOF^{WT–HA}$) to control for epitope-tag effects. Cells expressing $MYOF^{WT–FLAG}$ alone controlled for cross-reactivity of the anti-HA antibody, and cells expressing $MYOF^{WT–HA}$ alone controlled for non-specific binding to the beads; no HA signal was recovered in either immunoprecipitate. MYOF co-precipitated with itself in all combinations tested, indicating that MYOF self-associates and that $MYOF^{G1654S}$ retains the capacity to associate with wild-type MYOF. MYOF was detected at 230 kDa. Whole-cell lysates were immunoblotted with anti-HA and anti-FLAG antibodies, and band intensities were normalized to HSP90. Within each pair (**Fig. S4C** and **D**; **E** and **F**; **G** and **H**), immunoblots were derived from the same lysates. Spearman's correlation coefficient was used to test statistical significance. P value 0.05 was set as the significance threshold.

**Proximity ligation assay**

Interaction between MYOF and CaV1.2 was assessed using a commercially available proximity ligation assay (PLA) kit (Sigma Aldrich) per manufacturer's instructions. Briefly, iPSC-CMs were seeded on day 20 of differentiation in Lab-Tek II chamber slides (ThermoFischer, Cat# 154453) at a density of $2.5x10^5$ cells/well. On day 30, iPSC-CM were fixed with 4% paraformaldehyde (15 minutes) and permeabilized with 0.25% Triton X-100 (15 minutes). Specimens were blocked for 1 hour at 37 degrees C in a humidified chamber, followed by a primary antibody incubated overnight at 4 degrees C in antibody dilution buffer (1:50). Cells were incubated with PLA PLUS and MINUS probes for 1 hour and ligated for 3 minutes in a humidified chamber. The signal was amplified with polymerase overnight at 37 degrees C. Samples were washed and mounted with DAPI (Thermo Fischer, Cat# P36935). Images were acquired on an Olympus Fluoview 300 confocal microscope with a 60x oil immersion objective. Quantification was blinded and performed using ImageJ. Refer to **Table S3** for antibody information.

#### ***Myof*** $^{+/-}$ **mouse model generation**

The MYOF tm1a mice were generated by inserting loxP sites around the critical exons 6 and 7 (*19–21*). *Myof* $^{tm1a/tm1a}$ mice were crossed with the cardiac-specific alpha myosin-heavy chain (*Myh6*) promoter driver of Cre expression (αMyHC-Cre) (*22*) (**Fig. S7)**, generating heterozygous *Myof* $^{tm1b/+;\alpha MyHC\text{-}Cre}$ (*Myof* $^{+/-}$) were generated and validated for loss of MYOF protein expression in the cardiac tissue. *Myof* $^{tm1a/+}$ (*Myof* $^{+/+}$) mice were used as a control.

#### **Echocardiography**

Echocardiography was performed on 6-8-months old mice using a 15-45MHz transthoracic transducer and a VisualSonics Vevo770 system (Fujifilm). Averages of multiple M-mode images were obtained from conscious mice after hair removal. Data were analyzed in a blinded manner using VevoLab LV trace. Left ventricular (LV) end-systolic volume (LVESV) and end-diastolic volume (LVEDV) were measured from M-mode recordings taken at the mid-LV cavity in the short-axis, mid-papillary plane using the Vevo LAB short-axis LV Trace software. Stroke volume (SV) was determined by subtracting LVESV from LVEDV (SV = LVEDV – LVESV). Cardiac output was then calculated as the product of stroke volume and heart rate (CO = SV × heart rate). Left ventricular ejection fraction (LVEF) was computed as (100 × SV / LVEDV).

**Electrocardiogram**

Mice were sedated with 3% isoflurane in 100% $O_2$ for induction and 1.5% for maintenance. Once anesthesia was established, the ECG signal was recorded for 5 min using ADInstruments Power Lab and Bio Amp. All ECG signals were analyzed blinded to mouse genotype using LabChart Reader (ADInstruments, v8.1.8). QRS amplitude was measured by subtracting the maximum signal at the R peak from the minimum signal at the S peak. The QRS duration was measured by calculating the distance between the first deflection following the P wave and the halfway point between the S peak and the J wave (*23*). J wave amplitude was measured by subtracting the maximum J wave point from the isoelectric point. QT was measured according to previously published guidelines (*24*). QT was only measured on recordings with minimal background noise.

**H&E sample preparation**

The tissue samples were processed overnight on a Leica Peloris II tissue processor. The 12 hour processing schedule consisted of 1 station of 70% ETOH (diluted 100% ETOH), 2 stations of 95% ETOH (Cat# 2801, Decon Labs, King of Prussia, PA), 3 stations of 100% ETOH (Cat# 2701, Decon Labs, King of Prussia, PA), 3 stations of xylene (Cat# 3803665, Leica, Buffalo Grove, IL) and 3 stations of paraffin (Cat# 39602004, Paraplast Plus, Leica, Buffalo Grove, IL). The reagents with multiple stations contained diluted percentages in strength, with the last station in each being 100% pure.

Following processing, the samples were embedded in paraffin (Paraplast Plus, Leica, Buffalo Grove, IL). The tissue blocks were sectioned at 4µm utilizing a Leica RM2235 microtome. The sections were floated onto a 44°C waterbath and picked up, using Superfrost Plus slides (Cat# 12-550-15, Thermo Fisher Scientific, Waltham, MA). The slides were then baked at 60°C for 30 minutes. Following baking, the slides were loaded onto a Leica ST5020 with integrated coverslipper, CV5030, for Hematoxylin and Eosin (H&E) staining. The H&E staining reagents were the Leica SelecTech stain line, utilizing the Hematoxylin 560MX (Cat# 3801575), Blue Buffer (Cat#3802918), Aqua Define MX (Cat# 3803598), and Eosin 515 Phloxine (Cat# 3801606).

**PicroSirus Red sample preparation**

The Picro-Sirius Red staining technique was performed in accordance with a standard histologic staining protocol. The protocol consisted of placing slides in Weigert's Hematoxylin working solution for 5 minutes and thoroughly rinsing. The slides were then transferred to the Picro-Sirius Red staining solution for 60 minutes. The slides were placed in 0.5% Periodic Acid solution and differentiated in 2 changes for 5 seconds each or until collagen and fibrous tissue are red, surrounded by a yellow background, microscopically. The slides were then dehydrated in ascending grades of ETOH and cleared in xylene. From there, they were coverslipped using the Leica CV5030 mentioned above.

**Isolation of ventricular cardiomyocytes**

Ventricular cardiomyocytes were isolated from *Myof* $^{+/-}$ and *Myof* $^{+/+}$ mice using a Langendorff retrograde perfusion protocol as previously described (*25*). Briefly, anesthetized mice underwent cardiac excision followed by perfusion with modified Tyrode's solution containing Liberase (Roche) to achieve enzymatic extracellular matrix digestion. Ventricular tissue was then dissected away from the atria, and digestion was terminated in fetal bovine serum (10%). Isolated cardiomyocytes were resuspended in stop buffer, subjected to stepwise calcium re-introduction to 1 mM, and allowed to equilibrate for 1 hour before experimentation at room temperature.

**Patch clamp of ventricular cardiomyocytes**

Whole cell patch clamp recordings were conducted using a HEKA EPC10 using Patchmaster (v2x90.2) software (Harvard Bioscience, Inc, Holliston, MA). Data were acquired at 10-20 kHz. Borosilicate glass electrodes (World Precision Instruments, Sarasota, FL) pulled with a Sutter P-97 (Sutter Instruments, Novato, CA) to a resistance of 1-2 MΩ when filled with internal solution (see below). Amplifier circuitry was used for capacitance and series resistance compensation. Cells were held at -60 mV. A p/4 leak subtraction protocol was employed from a holding potential of −90 mV for all protocols.

A series of voltage-clamp protocols were used to characterize the biophysical properties of VGCCs in cardiac myocytes. The first of these was an I-V protocol, which consisted of 50 ms test pulses to potentials ranging between -80 and +50 mV in 10 mV steps, followed by a return step to -70 mV. This protocol was run with an inter-sweep interval (ISI) of 5 s. The second protocol was used to estimate steady-state inactivation. This protocol consisted of a 500 ms conditioning step ranging from -100 to +10 mV, in 10 mV increments, followed by a 10 ms step back to -80 mV (to enable open channels to close), which was followed by a 50 ms test pulse to the potential corresponding to the peak inward current (generally between 0 and +10 mV). The third protocol was used to assess recovery from inactivation. This protocol consisted of a 100 ms conditioning step to +10 mV to inactivate currents, followed by a voltage step to -60 mV for exponentially increasing duration to allow recovery from inactivation, and then a 30 ms test pulse to +10 mV. The longest recovery pulse duration was 1020 ms. The ISI for this protocol was also 5 s. Finally, to assess the voltage-dependence of channel deactivation, a tail current protocol was used consisting of a 10 ms conditioning step to +10 mV to activate currents, followed by a voltage step to potentials ranging from -100 to -20 mV. This protocol was run with an ISI of 5 s. Data were analyzed with Fitmaster V2x92.

*Solutions*

To isolate $Ca^{2+}$ currents, we used an extracellular solution containing: 100 mM Choline-Cl, 3 mM KCl, 10 mM HEPES, 10 mM glucose, 5 mM $CaCl_2 \cdot 2H2O$, 0.6 mM $MgCl_2$, adjusted to a pH 7.4 with Tris-Base and to 325 mOsm with sucrose. The electrode solution was composed of 100 mM CsCl, 40 mM TEA-Cl, 1 mM $CaCl_2$, 2 mM $MgCl_2$, 11 mM EGTA, 10 mM HEPES, 2 mM ATP-Mg, 1 mM GTP, adjusted to a pH 7.4 with Tris-Base and 310 mOSm with sucrose.

**Supplementary Tables**

**Table S1. Unrelated cases with cardiac involvement in MYOF p.G1654S carriers.**

Patients in Penn and BioVU biobanks were evaluated for phenotypes associated with MYOF p.G1654S. Age and sex were removed for patient privacy and are available upon request. Abbreviations: Penn (University of Pennsylvania Biobank); BioVU (Vanderbilt University Medical Center Biorepository); VT (ventricular tachycardia); SCD (sudden cardiac death); HF (heart failure); EF (ejection fraction); CAD (coronary artery disease); PVC (premature ventricular complexes).

| **ID** | **Cardiovascular History** |
|---|---|
| **Penn 1** | Non ischemic cardiomyopathy with heart failure and VT requiring heart transplant. Family history of SCD. |
| **BioVU 1** | Nonischemic VT. No known family history of HF or SCD. |
| **BioVU 2** | Stress cardiomyopathy (EF 40%) no CAD. Two years after event, recovered to 55%; PVCs on ECG and 5 beats of VT on heart monitor. |
| BioVU 3 | No documented cardiac history. |
| BioVU 4 | Left ventricular hypertrophy; no CAD. |
| BioVU 5 | No documented cardiac history. |

**Table S2. Description of sgRNAs and HDR repair templates used for CRISPR/Cas9. editing.** Spy Cas9 was used for rescue c.4960A>G and knock in c.4960G>A targeting NGG PAM, and SpRY Cas9 was used for knock in c.4723G > C targeting NNN PAM.

| iPSC line | sgRNA | HDR | Cas9 |
|---|---|---|---|
| Rescue c.4960A>G | CTCCTCTGGTATGCTGCAGT | TAGGCCTCCCCAGGTGCCAATGTCAGAGGCAGCAAAGTGCCTCTTGTACAGTCAAAGCAACTTACACACAGTACTCCTCTGGTATGCCGCAGTGGGACCCAAAGCGGGAAAGGAATCGGTTTTCCAGAT | Spy Cas9 NLS (NEB, Cat# M0646T) |
| Knock in c.4960G>A | CTCCTCTGGTATGCCGCAGT | TAGGCCTCCCCAGGTGCCAATGTCAGAGGCAGCAAAGTGCCTCTTGTACAGTCAAAGCAACTTACACACAGTACTCCTCTGGTATGCTGCAGTGAGACCCAAAGCGGGAAAGGAATCGGTTTTCCAGAT | Spy Cas9 NLS (NEB, Cat# M0646T) |
| Knock in c.4723G > C | TTTATGTAAGGGTCACACTG | GGTTGAGAGTGTTGGGAATGTAGTGATCTCGGTCTTCAATGACTTTTTGCCCAGTGTTATTTTATGTAAGGGTGACACTGTGGGACAAAATAGACGGGATGTTACATCATTGTAGGGCGCAAGGTGAAGTCAAGTTCAGGGACAGACTC | SpRY Cas9 (NEB, Cat# M0669T) |

**Table S3. Antibody information.**

| Antibody | Manufacturer | Catalogue # | Species | Dilution |
|---|---|---|---|---|
| **Immunoblot** | | | | |
| MYOF | QED Biosciences | 90020 | mouse | 1/1000 |
| GAPDH | Cell Signaling | D16H11 | rabbit | 1/1000 |
| JUP | ThermoFischer | 13-8500 | mouse | 1/1000 |
| Cav1.2 | Almone labs | ACC-003-GP | Guinea pig | 1/1000 |
| **Immunofluorescence** | | | | |
| CaV1.2 | US Biological Life Sciences | C2097-85B4 | mouse | 1/100 |
| Cav1.2 | Almone labs | ACC-003-GP | Guinea pig | 1/1000 |
| MYOF | Abcam | AB190264 | rabbit | 1/100 |
| JUP | ThermoFischer | A303-718A | rabbit | 1/100 |
| DSG2 | Santa Cruz | sc-08663 | mouse | 1/100 |
| Anti-α-Actinin (Sarcomeric) | Sigma | A7811 | mouse | 1/2500 |
| Anti-Cardiac Troponin T | Abcam | AB45932 | rabbit | 1/1000 |

| | | | | |
|---|---|---|---|---|
| SOX2 | Cell Signaling | L1D4A2 | mouse | 1/400 |
| OCT4A | Cell Signaling | C30A3 | rabbit | 1/400 |
| Nestin | Santa Cruz | sc-23927 | mouse | 1/1000 |
| Brachyury | Abcam | ab209665 | rabbit | 1/1000 |
| SOX17 | Cell Signaling | 81778S | rabbit | 1/1000 |
| **Proximity ligation assay** | | | | |
| CaV1.2 | US Biological Life Sciences | C2097-85B4 | mouse | 1/50 |
| MYOF | Abcam | AB190264 | rabbit | 1/50 |
| **Co-immunoprecipitation** | | | | |
| MYOF | QED Biosciences | 90020 | mouse | 1/1000 |
| Cav1.2 | Almone labs | ACC-003-GP | guinea pig | 1/1000 |
| HA | Cell Signaling | C29F4 | rabbit | 1/1000 |

**Table S4. Taqman primer information.**

| Taqman primer | Species | Assay ID | Catalogue number |
|---|---|---|---|
| MYOF | human | Hs00203853_m1 | 4331182 |
| GAPDH | human | Hs02786624_g1 | 4331182 |
| JUP | human | Hs00158408_m1 | 4331182 |
| Sox 2 | human | Hs04234836_s1 | 4331182 |
| c-Myc | human | Hs00153408_m1 | 4331182 |
| Oct-3/4 | human | Hs04260367_gh | 4331182 |
| Nanog | human | Hs07290356_m1 | 4331182 |

**Supplementary Figures**

I.1

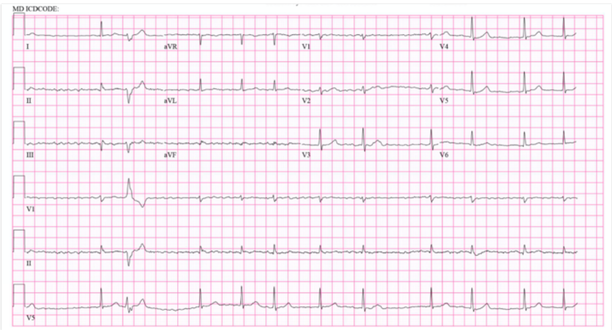

II.1

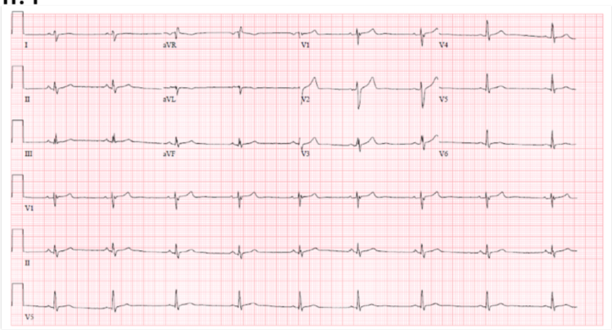

II.2

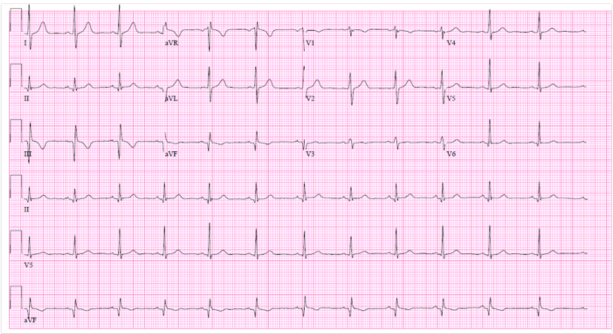

**Figure S1. Ambulatory ECG obtained from patients I.1, II.1, II.2.** I.1 showed rate-controlled atrial fibrillation with borderline nonspecific interventricular conduction delay and premature ventricular contraction. No T-wave inversions (TWI)**.** Signal from II.1 showed sinus bradycardia, borderline nonspecific interventricular conduction delay. No TWI. II.2 showed a normal sinus rhythm, early R wave transition with a prominent R wave in V1. No TWI.

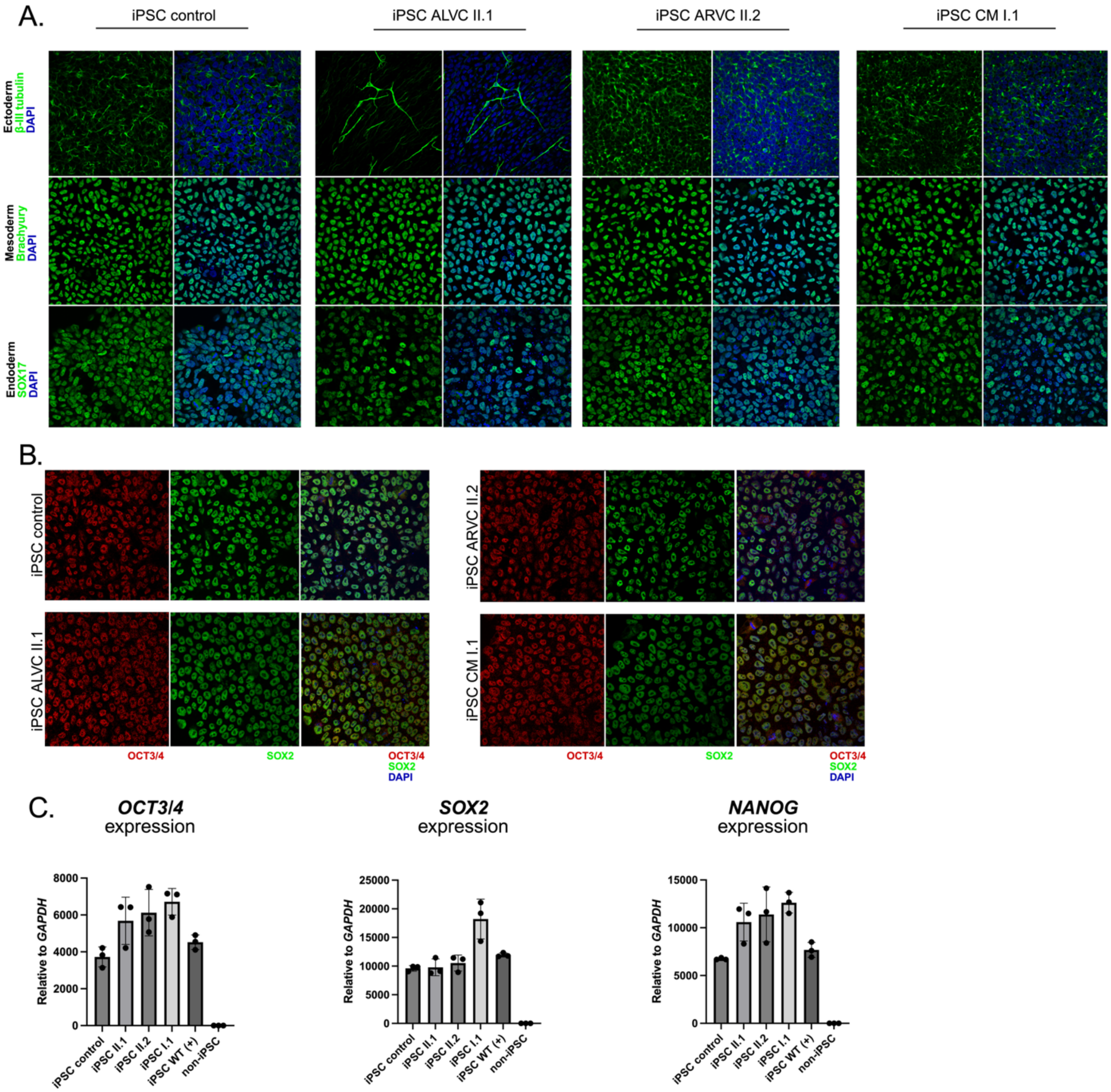


**Figure S2. Validation of pluripotency in iPSCs. A.** Three germ layer differentiation assay. Each iPSC line (iPSC control, ALVC II.1, ARVC II.2, CM I.1) was differentiated into ectoderm, mesoderm, or endoderm. Immunofluorescent staining for each representative marker is shown as follows: ectoderm, beta-III tubulin; mesoderm, brachyury; endoderm, SOX17. **B.** Immunofluorescent staining showing OCT3/4 and SOX2 expression in each iPSC line. **C.**

Transcript expression of *OCT3/4*, *SOX2*, and *NANOG* assessed by quantitative PCR. Newly established iPSC lines were compared to an established iPSC line in the lab (iPSC WT +) as a positive control and a non-iPSC cell type as a negative control.

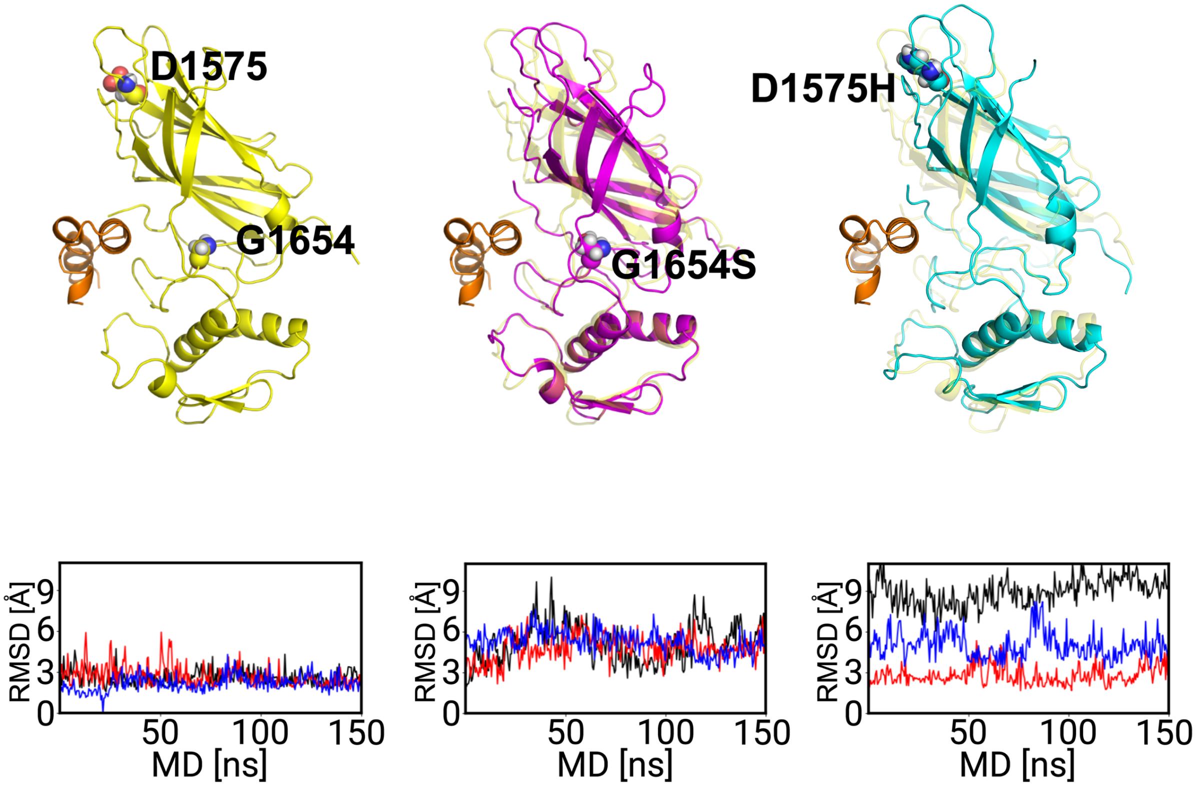


**Figure S3. Three independent molecular Dynamics (MD) simulations of C2F WT vs. p.G1654S vs. p.D1575H structures.** The linker domain is shown in orange to represent a change in its proximity to the C2 domain head. The C2F functional units were generated using AlphaFold. Minimal root-mean-squared-deviation (RMSD) (~1-3 Å) is shown in three independent simulations of the WT structure. C2F p.G1654S (magenta) overlaid with WT (yellow) C2F showing deviation of the mutant C2 domain head from the WT axis and increased RMSD (~3-6 Å). p.D1575H (cyan) overlaid with WT (yellow) showing increased RMSD (~3-9 Å). Variants are shown as spheres.

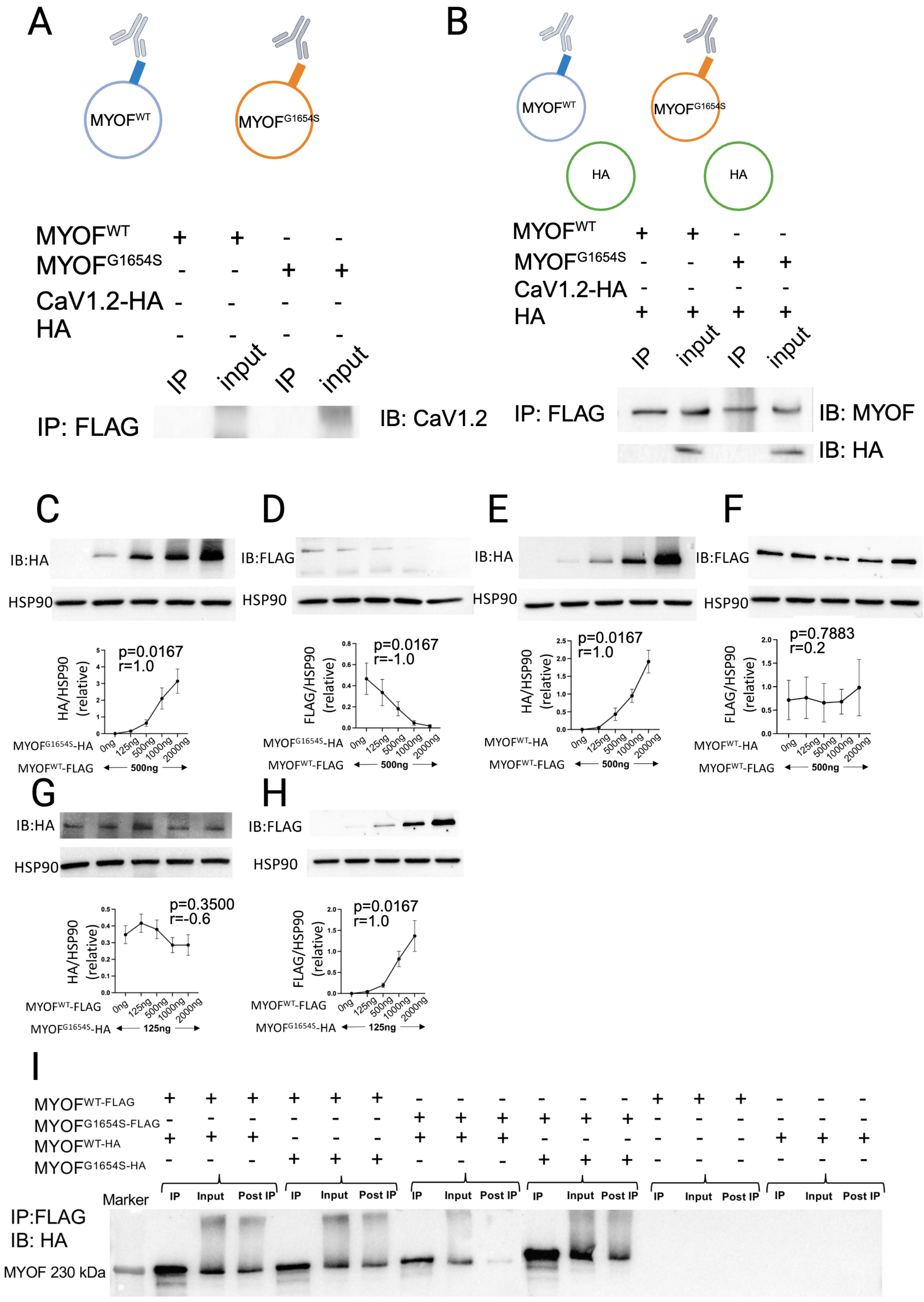
A
MYOFWT
MYOFG1654S
MYOFWT + + - -
MYOFG1654S - - + +
CaV1.2-HA - - - -
HA - - - -
IP input IP input
IP: FLAG
IB: CaV1.2
B
MYOFWT
MYOFG1654S
HA
HA
MYOFWT + + - -
MYOFG1654S - - + +
CaV1.2-HA - - - -
HA + + + +
IP input IP input
IP: FLAG
IB: MYOF
IB: HA
C
IB:HA
HSP90
HA/HSP90 (relative)
p=0.0167
r=1.0
MYOFG1654S-HA 0ng 125ng 500ng 1000ng 2000ng
MYOFWT-FLAG 500ng
D
IB:FLAG
HSP90
FLAG/HSP90 (relative)
p=0.0167
r=-1.0
MYOFG1654S-HA 0ng 125ng 500ng 1000ng 2000ng
MYOFWT-FLAG 500ng
E
IB:HA
HSP90
HA/HSP90 (relative)
p=0.0167
r=1.0
MYOFWT-HA 0ng 125ng 500ng 1000ng 2000ng
MYOFWT-FLAG 500ng
F
IB:FLAG
HSP90
FLAG/HSP90 (relative)
p=0.7883
r=0.2
MYOFWT-HA 0ng 125ng 500ng 1000ng 2000ng
MYOFWT-FLAG 500ng
G
IB:HA
HSP90
HA/HSP90 (relative)
p=0.3500
r=-0.6
MYOFWT-FLAG 0ng 125ng 500ng 1000ng 2000ng
MYOFG1654S-HA 125ng
H
IB:FLAG
HSP90
FLAG/HSP90 (relative)
p=0.0167
r=1.0
MYOFWT-FLAG 0ng 125ng 500ng 1000ng 2000ng
MYOFG1654S-HA 125ng
I
MYOFWT-FLAG + + + + + + - - - - - - + + + - - -
MYOFG1654S-FLAG - - - - - - + + + + + + - - - - - -
MYOFWT-HA + + + - - - + + + - - - - - - + + +
MYOFG1654S-HA - - - + + + - - - + + + - - - - - -
Marker
IP Input Post IP IP Input Post IP IP Input Post IP IP Input Post IP IP Input Post IP IP Input Post IP
IP:FLAG
IB: HA
MYOF 230 kDa

**Figure S4**. **Controls for the MYOF–CaV1.2 co-immunoprecipitation, dose-dependent depletion of MYOF$^{WT}$ by MYOF$^{G1654S}$, and MYOF self-association.** (**A** and **B**) Controls for the co-immunoprecipitation shown in **Fig. 6F–G**. HEK293T cells were transfected with MYOF$^{WT–FLAG}$ or MYOF$^{G1654S–FLAG}$ in the absence of the CaV1.2$^{HA}$ expression plasmid. (**A**) MYOF$^{WT}$ and MYOF$^{G1654S}$ were expressed without CaV1.2 and pulled down with anti-FLAG magnetic beads to control for non-specific binding and antibody detection; no CaV1.2 was detected. (**B**) MYOF$^{WT}$ and MYOF$^{G1654S}$ were co-expressed with a control vehicle plasmid (pcDNA3.1$^{HA}$) lacking the CaV1.2 insert to control for non-specific interaction with the epitope tag; the HA tag did not co-immunoprecipitate with MYOF. The experiment was repeated at least three times. (**C** to **H**) Dose-dependent depletion of MYOF$^{WT}$ by MYOF$^{G1654S}$. (**C** and **D**) A fixed amount of MYOF$^{WT–FLAG}$ (500 ng) was co-transfected with increasing amounts of MYOF$^{G1654S–HA}$ (0, 125, 500, 1000, and 2000 ng). (**C**) By anti-HA immunoblot, MYOF$^{G1654S–HA}$ accumulated in a dose-dependent fashion (p=0.0167, r=1.0). (**D**) By anti-FLAG immunoblot, MYOF$^{WT–FLAG}$ demonstrated progressive depletion with increasing MYOF$^{G1654S–HA}$ expression (p=0.0167, r=-1.0). (**E** and **F**) As a specificity control, an identical titration was performed with MYOF$^{WT–HA}$ (0–2000 ng) against a fixed amount of MYOF$^{WT–FLAG}$ (500 ng). (**E**) By anti-HA immunoblot, dose-dependent expression of MYOF$^{WT–HA}$ was shown (p=0.0167, r=1.0). (**F**) By anti-FLAG immunoblot, MYOF$^{WT–FLAG}$ levels were unchanged across the dose range (p=0.7883, r=0.2). (**G** and **H**) To investigate the effects of reciprocal titration, increasing amounts of MYOF$^{WT–FLAG}$ (0–2000 ng) were co-transfected with a fixed amount of MYOF$^{G1654S–HA}$ (125 ng). (**G**) By anti-HA immunoblot, MYOF$^{G1654S–HA}$ levels were not significantly altered by increasing wild-type protein (p=0.0.3500, r=-0.6). (**H**) By anti-FLAG immunoblot, MYOF$^{WT–FLAG}$ showed dose-dependent expression of (p=0.0167, r=1.0). For (**C**) to (**H**), representative immunoblots are shown; graphs are shown as mean ± SD from n=3 independent transfections for each sample. P and r values were computed using Spearman's correlation coefficient. (**I**) Self-association of MYOF as assessed by co-immunoprecipitation. The experiment was repeated at least three times. Immunoprecipitation (IP), immunoblot (IB). Band intensities from the MYOF titration panels were quantified by densitometry and normalized to the HSP90 loading control.

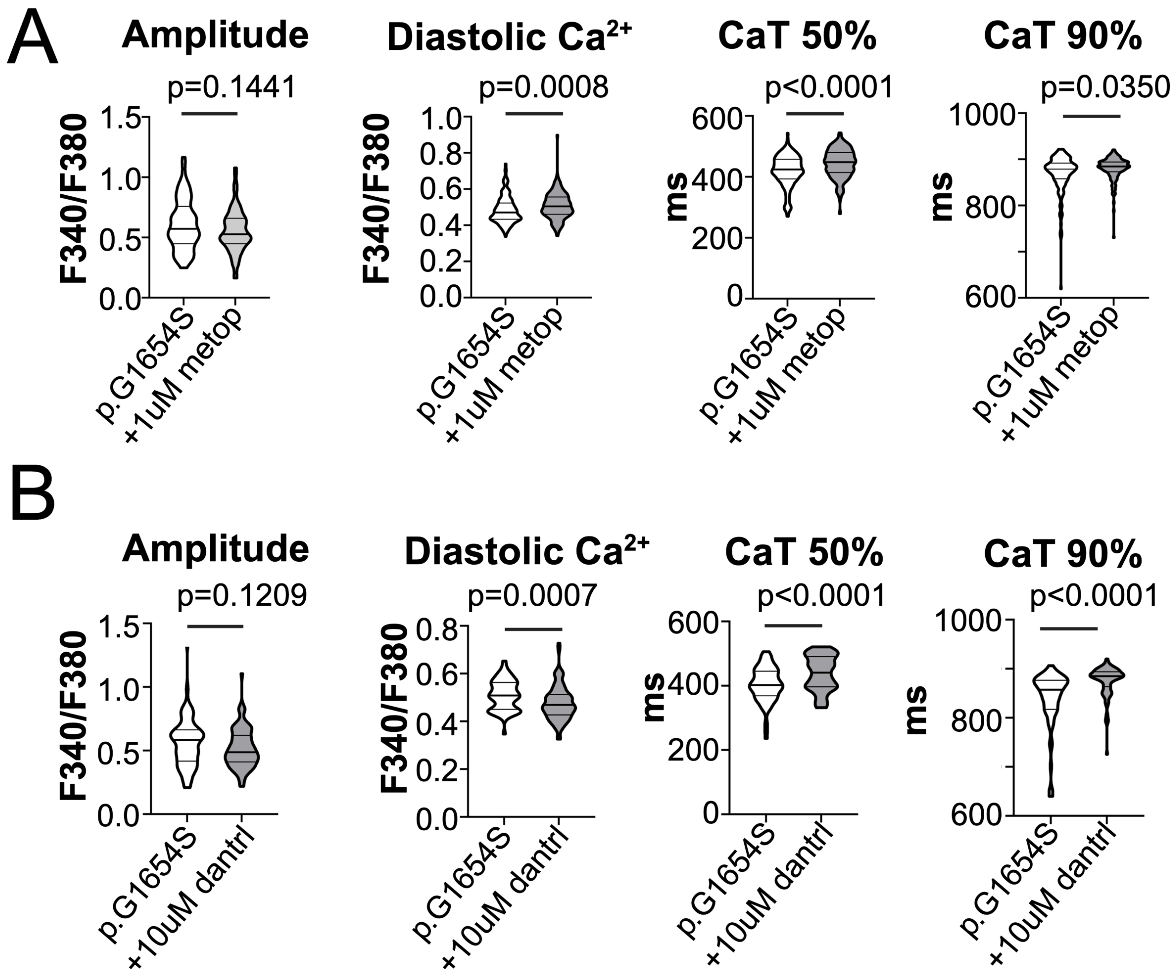


**Figure S5. Treatment with metoprolol or dantrolene did not rescue $Ca^{2+}$ handling defects.** **A.** Treatment with 1 µM metoprolol, a commonly used β-blocker in ACM management, worsened diastolic $Ca^{2+}$ overload and further prolonged CaT 50% and 90%. **B**. To determine whether $Ca^{2+}$ mishandling was due to SR leak via RYR2, we treated iPSC-CM with 10 µM dantrolene, a RYR2 inhibitor and stabilizer. Dantrolene treatment resulted in prolonged CaT at 50% and 90% and a modest reduction in diastolic $Ca^{2+}$ overload. Violin plots were reported as median ± IQR. Statistical testing was performed under the assumption that each cell was an independent unit of observation. Mann-Whitney test was used to assess statistical significance.

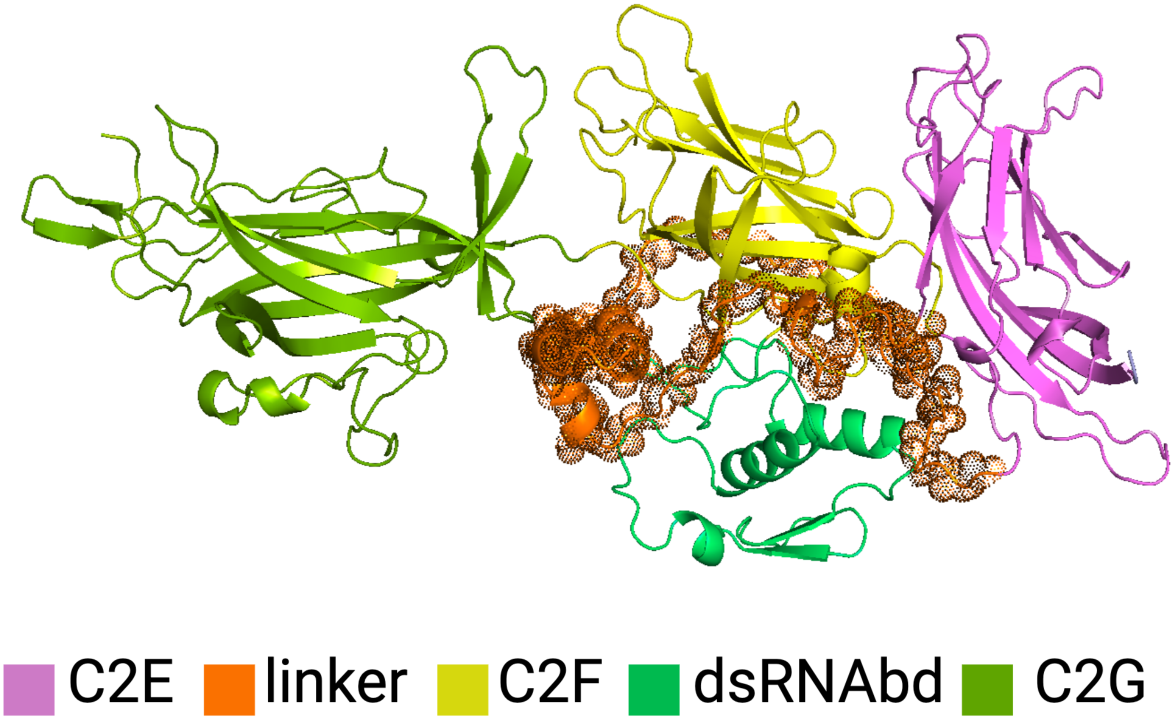


**Figure S6**. **AlphaFold modeling of the C2F domain.** AlphaFold predicted an unusual interaction feature, where a flexible linker encircles the C2F domain, creating a lever ring (‘linker domain’). Such a linker could provide a flexible substrate for docking with other membrane molecules or ion channels. Selection of C2G (1783-2008)-C2F (1545-1782)-linker (1406-1500)-double-stranded RNA binding domain (dsRNAbd) (1655-1740)-C2E (1301-1405; 1501-1545) functional units are shown and color coded.

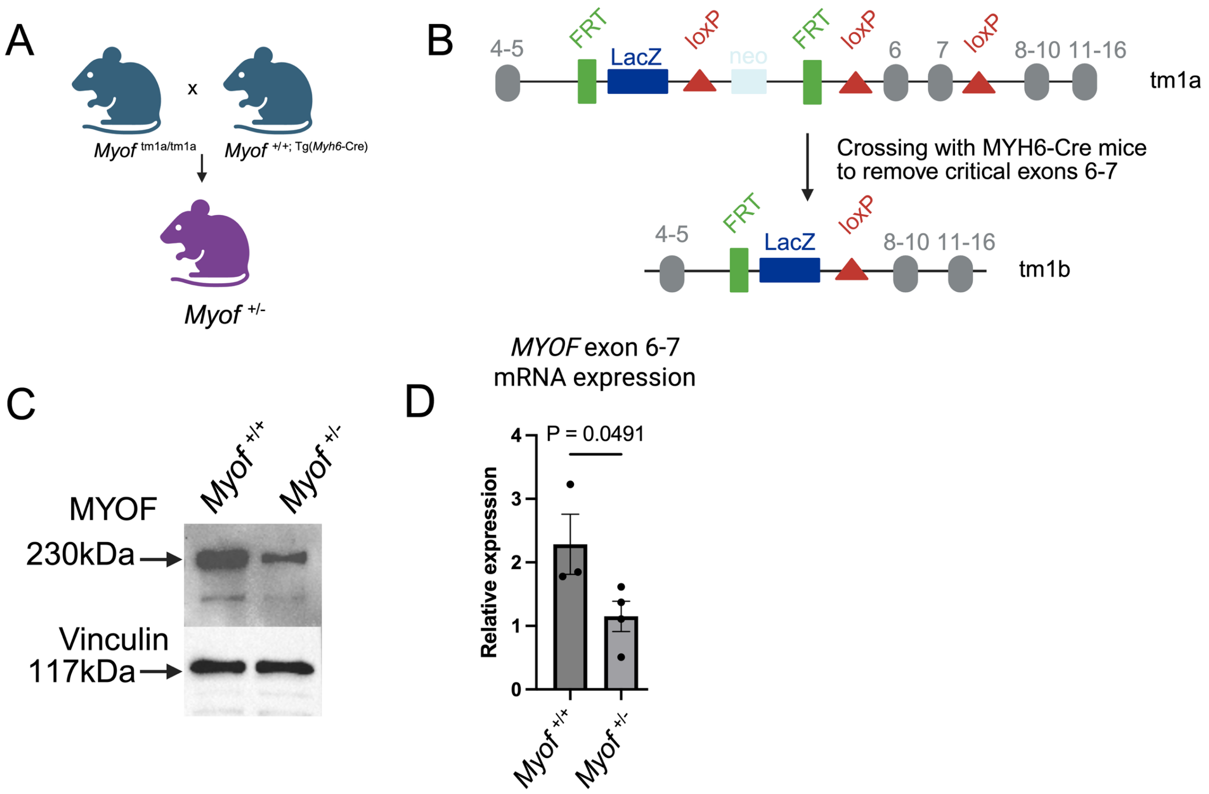


**Figure S7. Establishing *Myof* $^{+/-}$ mouse model and validating haploinsufficiency. A**. The homozygous tm1a allele carrying C57BL/6 male mice (*Myof* $^{tm1a/tm1a}$) were crossed with cardiac-specific αMyHC-Cre (*Myh6*-Cre) transgene carrying female with gene-matched background to generate cardiac-specific *Myof* $^{+/-}$. **B**. LoxP recombination cassette used to generate *Myof* $^{+/-}$. Tm1b mice (*Myof* $^{+/-}$) were generated as a result of recombination at loxP sites via the activity of *Myh6*-Cre. **C**. Validation of MYOF haploinsufficiency in *Myof* $^{+/-}$ heart tissue compared to *Myof* $^{+/+}$ controls. **D**. Decrease in mRNA expression of critical exons 6-7 in *Myof* $^{+/+}$ or *Myof* $^{+/-}$ heart tissue, confirming successful recombination and loss of critical exons. Data were shown as mean ± SD. P values were calculated by two-tailed unpaired Student's t-test.

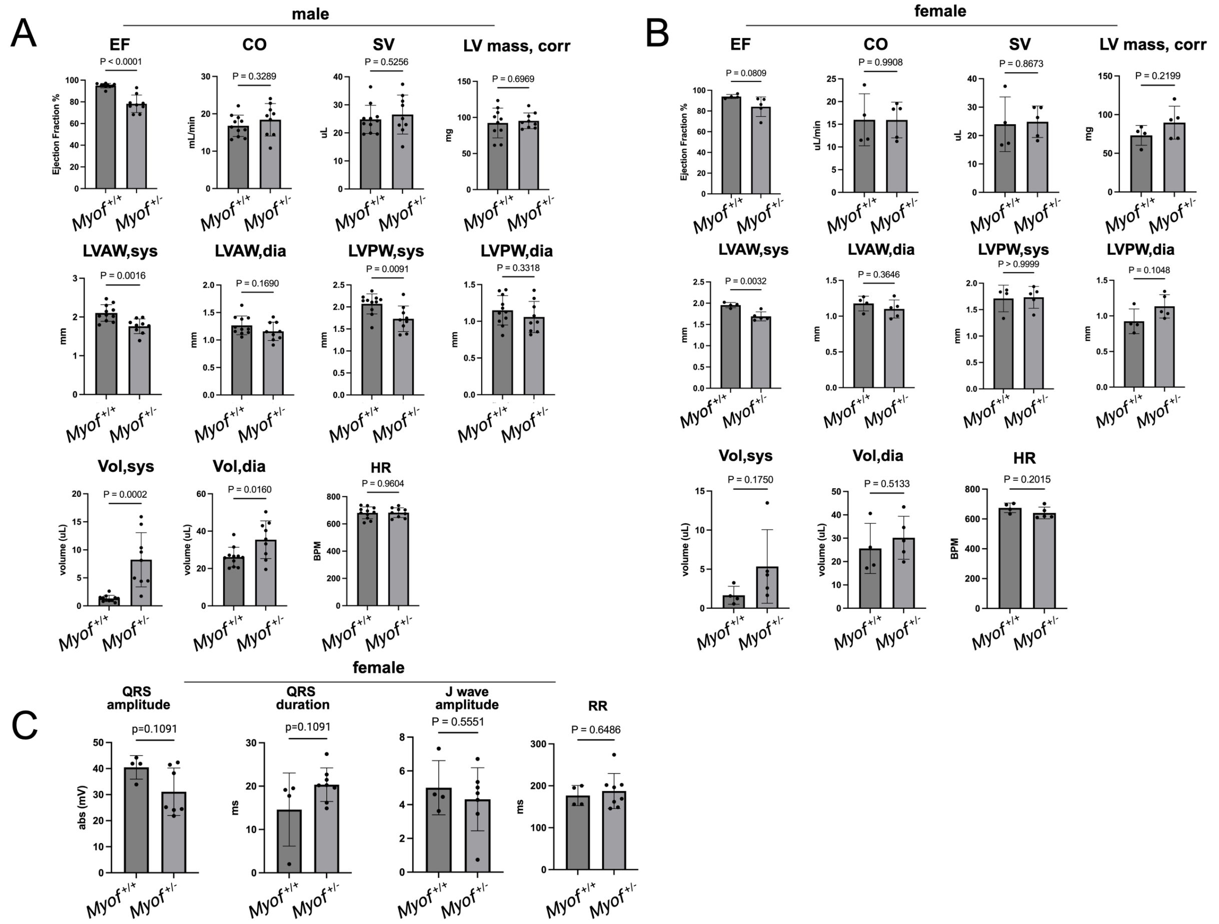


**Figure S8. Supplementary echocardiogram and electrocardiogram information for male or female *Myof* $^{+/-}$ vs. *Myof* $^{+/+}$ mice**. **A.** Supplementary echocardiogram information for *Myof* $^{+/-}$ male mice (n=11) compared to *Myof* $^{+/+}$ controls (n=9). Ejection fraction (EF), cardiac output (CO), stroke volume (SV), left ventricular mass corrected (LV mass, corr); left ventricular anterior wall, systolic (LVAW, sys); left ventricular anterior wall, diastolic (LVAW, dia); left ventricular posterior wall, systolic (LVPW, sys); left ventricular anterior wall, diastolic (LVAW, dia); volume, systolic (Vol, sys); volume, diastolic (Vol, dia). **B.** Supplementary echocardiogram information for *Myof* $^{+/-}$ female mice (n=6) compared to *Myof* $^{+/+}$ controls (n=4). **C.** Summary of electrical abnormalities in female *Myof* $^{+/-}$ mice (n=7) compared to *Myof* $^{+/+}$ controls (n=4). Quantification of ECG parameters. QRS amplitude, QRS duration, J wave amplitude, RR interval. QT was not quantified due to excessive background noise. Data are shown as mean ± SD. P values were calculated by two-tailed unpaired Student's t-test. Mann-Whitney test was used in (**A**, EF; **B**, LVPW, sys; **C**, QRS amplitude and duration) due to non-normal distribution.

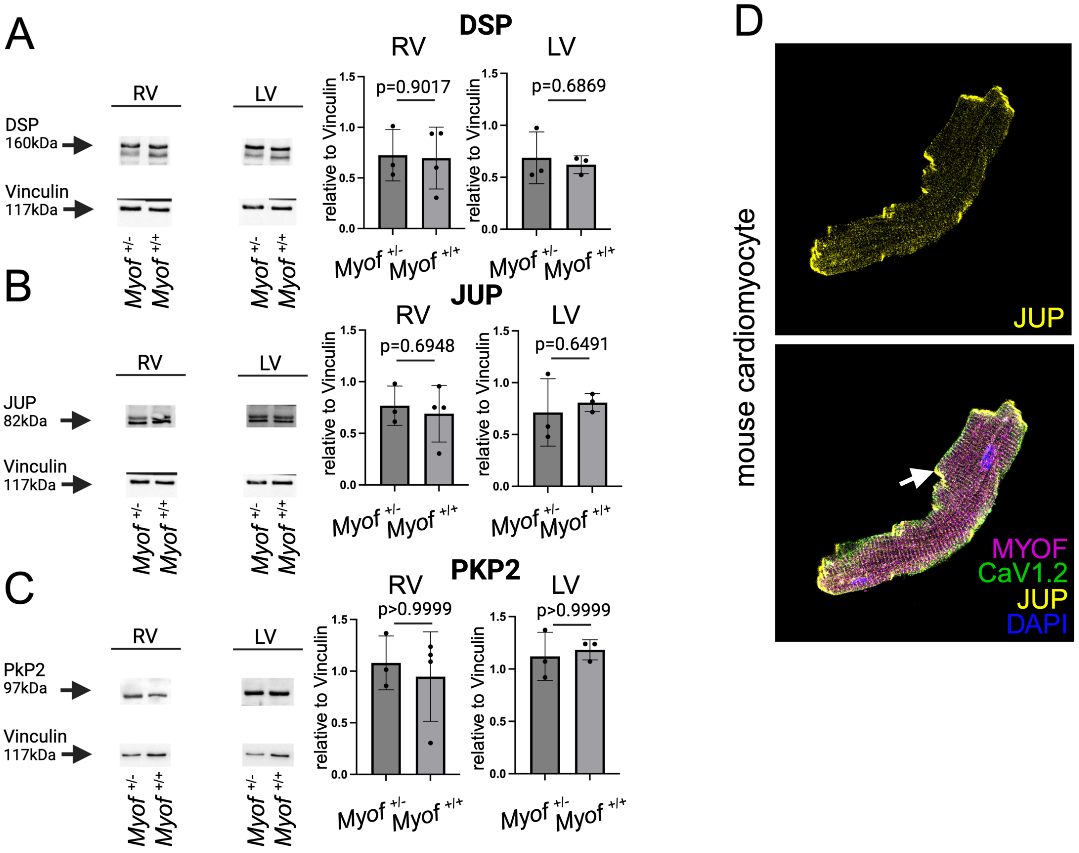


**Figure S9. Expression of desmosomal proteins in RV and LV tissue. A**. DSP expression unchanged in *Myof* $^{+/-}$ vs. *Myof* $^{+/+}$ heart tissue. **B**. JUP expression unchanged in *Myof* $^{+/-}$ vs. *Myof* $^{+/+}$ heart tissue. **C**. PKP2 expression unchanged in *Myof* $^{+/-}$ vs. *Myof* $^{+/+}$ heart tissue. The following samples were used from *Myof* $^{+/-}$ mice (RV n=3, LV n=3) compared to *Myof* $^{+/+}$ controls (RV n=4, LV n=3). **D**. Dissociated adult mouse cardiomyocytes. MYOF was co-stained with CaV1.2 and JUP. Overlayed images showing co-localization or lack thereof. MYOF co-localized with CaV1.2. Poor co-localization of JUP desmosomal components with MYOF. Data were shown as mean ± SD. (**A**, **B**) P values were calculated by two-tailed unpaired Student's t-test. (**C**) Mann-Whitney test was used due to non-normal distribution.

5. J. Cadrin-Tourigny, L. P. Bosman, A. Nozza, W. Wang, R. Tadros, A. Bhonsale, M. Bourfiss, A. Fortier, Ø. H. Lie, A. M. Saguner, A. Svensson, A. Andorin, C. Tichnell, B. Murray, K. Zeppenfeld, M. P. van den Berg, F. W. Asselbergs, A. A. M. Wilde, A. D. Krahn, M. Talajic, L. Rivard, S. Chelko, S. L. Zimmerman, I. R. Kamel, J. E. Crosson, D. P. Judge, S. C. Yap, J. F. van der Heijden, H. Tandri, J. D. H. Jongbloed, M. C. Guertin, J. P. van Tintelen, P. G. Platonov, F. Duru, K. H. Haugaa, P. Khairy, R. N. W. Hauer, H. Calkins, A. S. J. M. Te Riele, C. A. James, A new prediction model for ventricular

arrhythmias in arrhythmogenic right ventricular cardiomyopathy. *Eur. Heart J.* **43**, e1–e9 (2022).

18. S. Watkins, *Curr. Protoc. Cytom.*, in press, doi:10.1002/0471142956.cy1215s48.
